# Central Nervous System Complications in Miliary Pulmonary Tuberculosis: A Systematic Review Of Published Case Reports And Case Series

**DOI:** 10.1101/2024.07.07.24310052

**Authors:** Ravindra Kumar Garg, Vimal Kumar Paliwal, Anand Srivastava, Rajiv Garg, Sandeep Bhattacharya, Amita Jain, Imran Rizvi

## Abstract

**Background:** This systematic review aims to evaluate central nervous system involvement in context of miliary pulmonary tuberculosis.

**Methods:** We included studies on central nervous system infections associated with miliary pulmonary tuberculosis involving human subjects in any language. Patients with disseminated tuberculosis involving two non-contiguous organs were excluded. Databases searched included PubMed, Scopus, Embase, and Google Scholar. Case reports, case series, and observational studies with confirmed diagnoses were included. The review follows PRISMA guidelines and is registered with PROSPERO (CRD42024542130).

**Results:** The review included 158 records describing 161 patients. The mean age was 32.7 years, with 53.4% females. Key comorbid conditions included human immunodeficiency virus positivity (6.8%), immunosuppressive therapy (9.9%), and chronic kidney disease (2.5%). Common clinical features were fever (58.4%), headache or vomiting (41.6%), and altered sensorium (26.7%). Frequent neuroimaging abnormalities were miliary or multiple brain tuberculomas (66.5%), basal exudates (7.5%), vertebral tuberculosis (7.5%), cerebral infarcts (6.8%), and spinal cord tuberculoma or abscess (6.2%). Paradoxical manifestations occurred in one-third of cases, including new or enlarged brain lesions (22.4%) and paradoxical cerebral infarcts (2.5%). Cerebrospinal fluid findings were normal or unavailable in 45.3% of cases. Positive diagnostic tests included smear, culture, polymerase chain reaction, or GeneXpert in 25% of cases, with 5% showing drug resistance. Improvement was seen in 80.8% of patients, while 9.9% either died or did not improve.

**Conclusion:** This review highlights diverse central nervous system involvement in miliary pulmonary tuberculosis, including tuberculous meningitis and spinal cord lesions, with common paradoxical manifestations and significant patient improvement with following treatment.

## Introduction

Miliary tuberculosis is a severe and potentially life-threatening form of the disease characterized by the widespread dissemination of *Mycobacterium tuberculosis* throughout the body. This condition typically arises from the hematogenous spread of tubercle bacilli from the lungs to various organs via the bloodstream. The term “miliary” derives from the Latin word “miliarius,” referring to the millet seed-sized tubercular foci that develop in affected tissues.^1,2^ (Figure-1)

**Figure-1.**
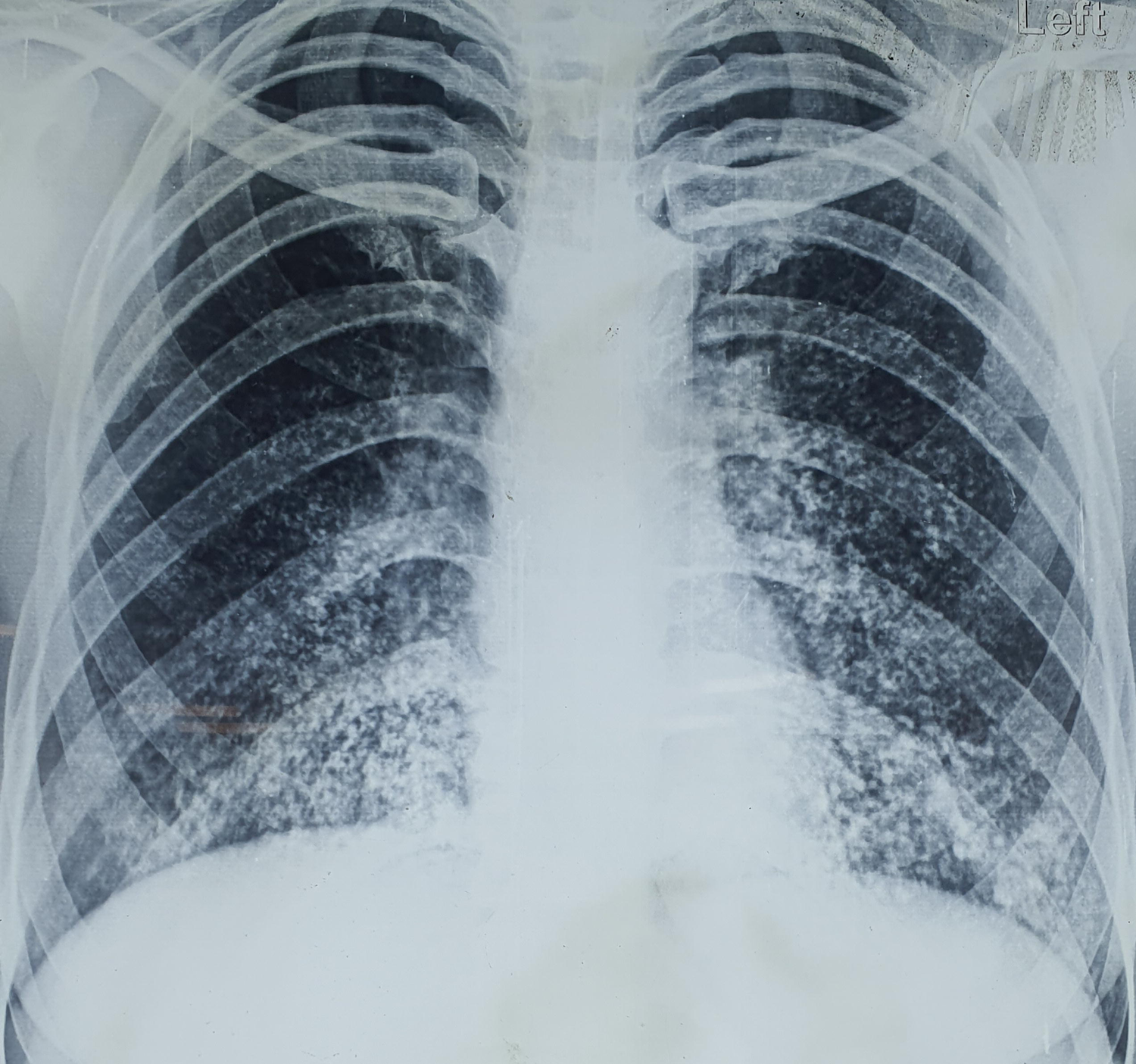
X-ray chest shows miliary tuberculous lesions of the lungs.

When *Mycobacterium tuberculosis* disseminates hematogenously throughout the body, it can potentially reach the central nervous system, leading to various neurological complications. The pathogenesis of central nervous system involvement in miliary tuberculosis typically occurs through the hematogenous spread of tubercle bacilli, which can breach the blood-brain barrier and infect the brain and its surrounding structures. Once within the central nervous system, *Mycobacterium tuberculosis* can cause a spectrum of neurological manifestations, including meningitis, tuberculomas, abscesses, and spinal cord involvement.^3^

This systematic review aims to identify and analyze the types, frequency, and clinical significance of central nervous system manifestations in patients with pulmonary miliary tuberculosis by examining case reports and series, providing a comprehensive understanding of the neurological complications associated with this severe tuberculosis form.

## Methods

This systematic review complies with the guidelines outlined in the PRISMA (Preferred Reporting Items for Systematic Reviews and Meta-Analyses) statement, as specified in the PRISMA checklist. Our review has been registered with PROSPERO (registration number: CRD42024542130).^4^ Approval from the Institutional Ethics Committee was not required for this systematic review, and no human subjects were enrolled.

### Inclusion and Exclusion Criteria

This systematic review included studies focusing on CNS infections in the context of pulmonary miliary tuberculosis. Studies were required to be available in any language, involve human subjects, and provide information on the spectrum of CNS manifestations associated with miliary pulmonary tuberculosis. Excluded were studies not specifically addressing CNS infections in the context of miliary pulmonary tuberculosis, as well as animal studies, in vitro studies, conference abstract and review articles. We did not consider patients with disseminated tuberculosis. Disseminated tuberculosis was defined if there were involvement of two non-contiguous organs.^5^

### Literature Search Strategy

The databases of PubMed, Scopus, Embase, and Google Scholar were systematically searched for relevant literature. In Google Scholar, the search extended through the first 50 pages of results. No language restrictions were applied, and non-English articles were translated using Google Translate. The search terms included (“CNS tuberculosis” OR “central nervous system tuberculosis” OR “tuberculous meningitis” OR “TB meningitis” OR “neurotuberculosis” OR “intracranial tuberculoma” OR “tuberculoma” OR “tuberculous encephalopathy”) AND (“miliary tuberculosis” OR “disseminated tuberculosis”). The most recent search was done on 06 May 2024.

### Study Selection Process

This review included case reports, case series, and observational studies that provided individual patient details and confirmed diagnoses using cerebrospinal fluid through serology, genetic testing, or smear analysis. If histopathological examinations following brain biopsy or autopsy were performed, these were also recorded.

### Data Collection Method

Essential information from selected studies, including author details, publication year, patient demographics, diagnostic methods, treatment approaches, and outcomes related to central nervous system manifestations in patients with miliary pulmonary tuberculosis, was systematically extracted using a data extraction form.

### Definitions

Miliary lesions in the brain were defined if neuroimaging revealed multiple/ numerous small, ring, or nodular-enhancing lesions, each measuring less than 3 mm in diameter. ^6^(Figure-2) Paradoxical manifestations were defined in individuals with miliary tuberculosis as the enlargement of existing tuberculous brain lesions or the appearance of new brain lesions, despite an initial positive response to anti-tuberculosis therapy.^7^

**Figure-2.**
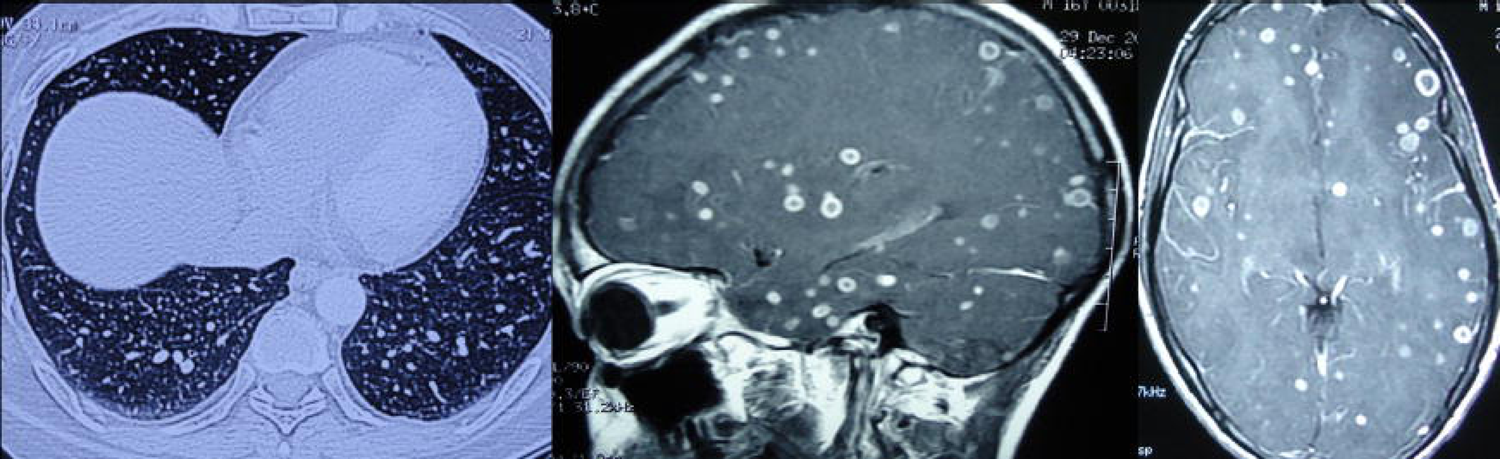
Magnetic resonance imaging of thorax and brain shows miliary tuberculous lesions of the lungs and brain.

### Data extraction

Two independent researchers (RK and VP) screened the titles and abstracts of search results to identify studies that met the criteria for CNS infections in miliary pulmonary tuberculosis. Full texts of potentially relevant studies were then thoroughly assessed. Any discrepancies in study inclusion were resolved through discussion or by consulting a third reviewer (AS). In cases of differing opinions, consensus was reached through discussion.

EndNote 21 software (Clarivate, Philadelphia, PA, USA) was used to manage duplicate entries. Any discrepancies or issues were resolved through consensus. A PRISMA flow diagram was prepared, detailing the number of records obtained and assessed at each stage, with the aid of the EndNote 21 tool.

### Quality assessment

Each case was evaluated based on four key criteria: selection, ascertainment, causality, and reporting, as outlined by Murad MH et al.^8^ According to Della Gatta et al.’s standards, a case report meeting all these criteria was deemed “good quality,” meeting three criteria was rated “fair quality,” and meeting only one or two criteria was considered “poor quality.”^9^

### Data Analysis

Demographic details, clinical features, comorbid conditions including HIV status, types and numbers of brain lesions, diagnostic procedures, final diagnoses, treatments, and patient outcomes were recorded in a Microsoft Word file. The consolidated data was then presented in tables, displaying the corresponding numbers and percentages.

## Results

Our review included 158 records describing 161 patients. (Supplementary item 1) Figure 3 presents the PRISMA flowchart for our systematic review. Of the 161 cases evaluated, 147 (91.3%) were classified as having good or fair quality, while the remaining 14 cases were deemed low quality (Supplementary Item 2). The PRISMA checklist is provided as Supplementary Item 3.

**Figure-3.**
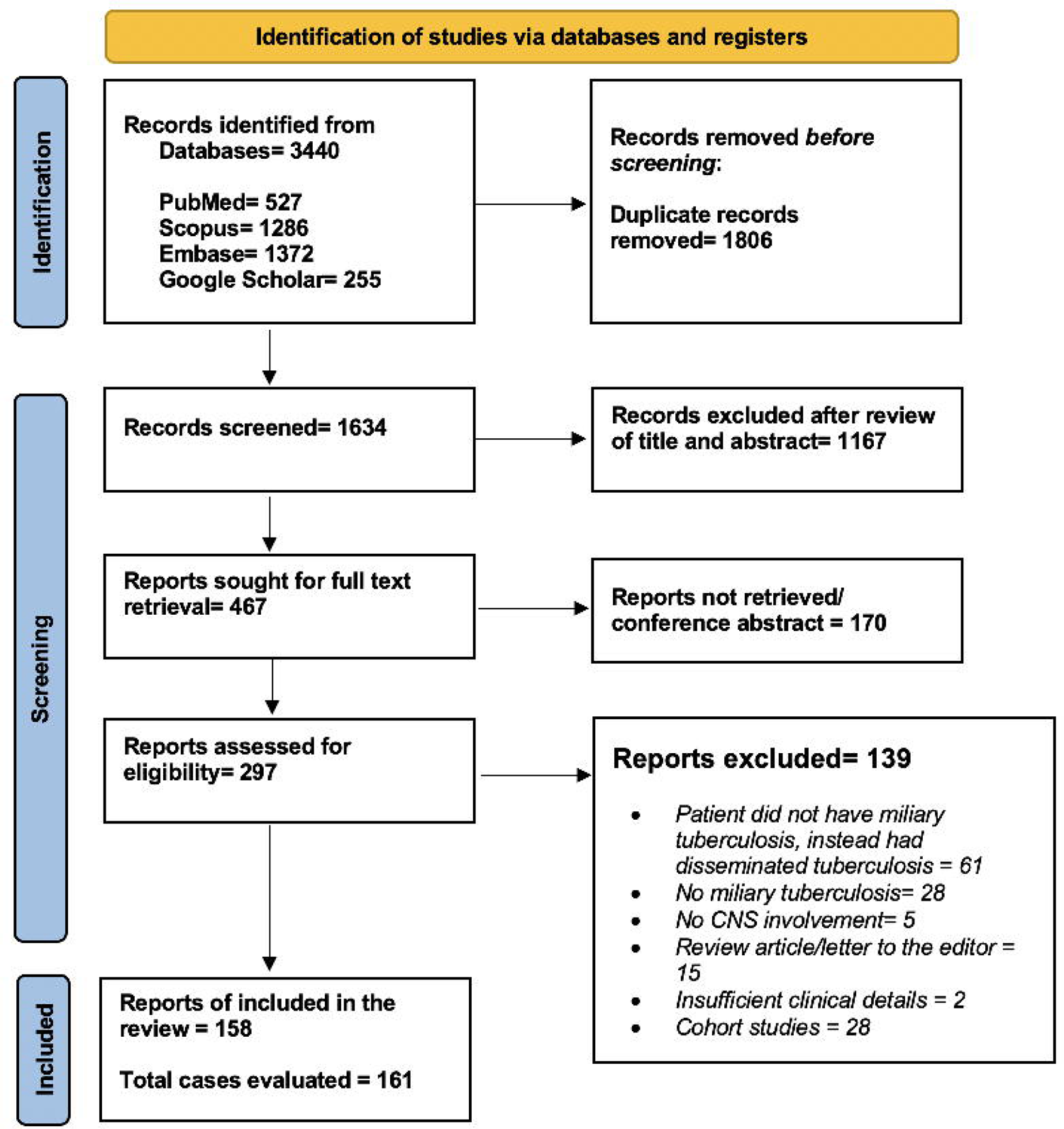
PRISMA flow diagram of the study depicts the procedure of selecting articles for the systematic review.

In this systematic review, the mean age was 32.7 years (median 31; range 1-91 years), including 11 infants. There were a total of 86 females (53.4%) and 73 males (45.3%). There were 15 immigrants (9.3%) and 5 pregnant ladies (3.1%). Our review found that 42 individuals had some comorbid condition. Among these, 11 patients (6.8%) were HIV positive, 16 (9.9%) were on immunosuppressive therapy, and 4 (2.5%) had chronic kidney disease (Table 1).

**Table-1:** Summary Of Clinical Characteristics of CNS Manifestations in Miliary Pulmonary Tuberculosis Patients (n=161)

| **Variables** | **Miliary Tuberculosis with CNS involvement (n=161)** |
| --- | --- |
| **Age (in years)** | NA= 1 (0.6%) |
|  | Less than 1 year= 11 (6.8%)<br>Mean= 32.7<br>Median = 31<br>Mode= 31<br>Range= 1-91<br>Interquartile<br>Range IQR= 27 |
| **Sex** | NA= 2 (1.2%)<br>M= 73 (45.3%)<br>F= 86 (53.4%) |
| **Other information** | Immigrants = 15 (9.3%)<br>Pregnant = 5 (3.1%) |
| **Co-morbid conditions** | NA/Negative = 119 (73.9%)<br>HIV positive= 11 (6.8%)<br>On immunosuppressive therapy= 16 (9.9%)<br>Renal transplant = 2 (1.2%)<br>Chronic kidney disease= 4 (2.5%)<br>COVID-19= 2 (1.2%)<br>Malnourishment= 2 (1.2%)<br>In vitro fertilization= 2 (1.2%)<br>Chronic liver disease= 2 (1.2%)<br>Noonan syndrome= 1 (0.6%) |
| **Duration of illness** | NA= 40 (24.8%)<br>Sudden= 5 (3.1%)<br>One week or less= 25 (15.5%)<br>One week up to one month= 54 (33.5%)<br>One month up 6 months= 36 (22.4%)<br>>6 months= 1 (0.6%) |
| **Common presenting clinical features** | Fever = 94 (58.4%)<br>Headache and/or vomiting = 67 (41.6%)<br>Altered sensorium = 43 (26.7%)<br>Focal neurological deficits = 30 (18.6%)<br>Seizures = 23 (14.3%)<br>Vision loss = 14 (8.7%)<br>Paraparesis/quadriparesis = 13 (8%)<br>Diplopia/Ophthalmoplegia = 7 (4.4%) |
| **Imaging abnormalities** | NA/Normal = 21 (13%)<br>Miliary/multiple tuberculoma of the brain =107 (66.5%)<br>Brain tuberculous abscess = 5 (3.1%)<br>Basal exudates =12 (7.5%)<br>Cerebral infarcts =11 (6.8%)<br>Spinal cord tuberculoma/abscess = 10 (6.2%)<br>Vertebral tuberculosis =12 (7.5%) |
| **Paradoxical CNS involvement** | NA/not reported = 111 (68.9%)<br>Paradoxical manifestations reported = 49 (30.4%)<br>Optochiasmatic arachnoiditis leading to vision loss = 3 (1.9%)<br>New brain lesions appeared or existing lesions enlarged = 36 (22.4%) |
|  | Paradoxical tuberculous meningitis =2 (1.2%)<br>Paradoxical cerebral infarcts = 4 (2.5%)<br>Paradoxical spinal cord lesions = 4 (2.5%)<br>Paradoxical vertebral lesions = 2 (1.2%)<br>Lymph adenopathy = 1 (0.6%) |
| **Eye tuberculous lesions** | Choroidal tubercles = 19 (11.8%)<br>Retinal lesions = 4 (2.5%) |
| **CSF abnormality** | NA/Normal = 73 (45.3%)<br>Abnormal = 88 (54.7%)<br>Smear/Culture/PCR/GeneXpert positive = 40 (25%) |
| **Drug resistance** | Present = 8 (5%)<br>MDR = 4<br>Mono-resistance = 2<br>Isoniazid and streptomycin resistance = 2 |
| **Biopsy/autopsy** | Brain biopsy = 14 (8.7%)<br>Vertebral/spinal biopsy = 5 (3.1%)<br>Lung biopsy = 4 (2.5%)<br>Lymph node biopsy = 7 (4.4%)<br>Other tissue biopsy = 11 (6.8%)<br>Autopsy = 5 (3.1%) |
| **Outcome** | NA = 15 (9.3%)<br>Died/not improved =16 (9.9%)<br>Improved = 130 (80.8%) |

CNS= Central Nervous System; F= Female; HIV= Human Immunodeficiency Virus; IQR= Interquartile Range; M= Male; MDR= Multidrug resistant; NA= Not Available; PCR= polymerase chain reaction

The most common presenting clinical features were fever in 94 individuals (58.4%), headache and/or vomiting in 67 (41.6%), and altered sensorium in 43 (26.7%). Other observed features included focal neurological deficits in 30 (18.6%), seizures in 23 (14.3%), vision loss in 14 (8.7%), paraparesis/quadriparesis in 13 (8%), and diplopia/ophthalmoplegia in 7 (4.4%). Choroidal tubercles were observed in 19 cases (11.8%), and retinal lesions were found in 4 cases (2.5%) (Table 1)..

The frequent neuroimaging abnormalities observed were miliary/multiple tuberculoma of the brain in 107 cases (66.5%), basal exudates in 12 cases (7.5%), vertebral tuberculosis in 12 cases (7.5%), cerebral infarcts in 11 cases (6.8%), spinal cord tuberculoma/abscess in 10 cases (6.2%), and brain tuberculous abscess in 5 cases (3.1%) (Table 1).

Paradoxical manifestations were not reported or were unavailable for 111 cases (68.9%). Among the reported cases (49 cases, 30.4%), new brain lesions appeared or existing lesions enlarged in 36 cases (22.4%), paradoxical cerebral infarcts occurred in 4 cases (2.5%), and paradoxical spinal cord lesions were observed in 4 cases (2.5%). Additionally, optochiasmatic arachnoiditis leading to vision loss was reported in 3 cases (1.9%), paradoxical tuberculous meningitis in 2 cases (1.2%), paradoxical vertebral lesions in 2 cases (1.2%), and lymphadenopathy in 1 case (0.6%) (Table 1).

In this review, cerebrospinal fluid findings were normal or unavailable in 73 cases (45.3%), while 88 cases (54.7%) showed abnormalities. Smear, culture, PCR, or GeneXpert tests were positive in 40 cases (25%), with 8 cases (5%) showing drug resistance, including 4 with multidrug resistance (MDR).

Brain biopsies demonstrating tuberculous lesions and the presence of *Mycobacterium tuberculosis* were performed in 14 cases (8.7%). Vertebral/spinal biopsies were done in 5 cases (3.1%), lung biopsies in 4 cases (2.5%), lymph node biopsies in 7 cases (4.4%), and other tissue biopsies in 11 cases (6.8%). Autopsies were conducted in 5 cases (3.1%). In our review, 16 patients (9.9%) died or did not improve, while 130 patients (80.8%) showed improvement.

## Discussion

In this systematic review, central nervous system involvement patterns included tuberculous meningitis, miliary tuberculoma brain, large tuberculomas, spinal cord lesions, intramedullary, intradural extramedullary tuberculomas, and tuberculous abscesses. Fever, headache/vomiting, and altered sensorium were the most common clinical features. Miliary/multiple brain tuberculomas were frequent neuroimaging findings. Paradoxical manifestations occurred in one-third of cases. Improvement was seen in majority of patients.

Several cohort studies on central nervous system involvement in miliary pulmonary tuberculosis have noted similar observations. For example, a cohort study assessed central nervous system involvement in miliary tuberculosis. Among 282 patients evaluated, 87.59% had central nervous system involvement. Magnetic resonance imaging showed the highest sensitivity (93.15 to 96.05%), outperforming cerebrospinal fluid examination (71.92 percent) and computed tomography (34.69%). Despite 59.65 percent bacteriological evidence from sputum, cerebrospinal fluid examination positivity was only 8.82 percent.^10^ Another prospective study from India found central nervous system involvement in 40% of 53 immunocompetent adults with pulmonary miliary tuberculosis, with 12 diagnosed with definite tuberculous meningitis.^11^ On the contrary, among 81 newly diagnosed tuberculous meningitis patients revealed thoracic computed tomography abnormalities in 69.1%, with centrilobular nodules being most common.^12^

Tuberculomas were the most common brain lesions, with magnetic resonance imaging abnormalities even in patients with normal cerebrospinal fluid, highlighting the need for systematic brain imaging. A study of 60 miliary tuberculosis patients with neurological symptoms found 80% had tuberculous meningitis and intracerebral tuberculomas in 27 patients.^3^ A retrospective analysis of 34 miliary tuberculosis patients showed cerebral involvement in over 60%, despite 13 having no neurological symptoms. Magnetic resonance imaging studies on seven patients with typical miliary tuberculosis but no central nervous system symptoms found brain involvement in all cases, reinforcing that brain involvement is common and imaging is vital for diagnosis and monitoring.^13^

Paradoxical central nervous system manifestations in miliary pulmonary tuberculosis frequently occur when patients experience a worsening of symptoms or the emergence of new lesions during antituberculous treatment. This phenomenon, often attributed to immune reconstitution inflammatory syndrome, happens as the immune system begins to recover and aggressively responds to residual mycobacterial antigens. This exaggerated immune response can lead to increased inflammation, expansion of existing tuberculomas, or formation of new lesions in the brain and meninges. These paradoxical reactions complicate the clinical course, necessitating careful monitoring and sometimes the use of corticosteroids to manage the inflammatory response and prevent further neurological damage.^7^ Paradoxical reaction and immune reconstitution inflammatory syndrome in tuberculosis can in refractory cases be treated with TNF-α antagonists after corticosteroids.^14^

We suggest an updated theory for the pathogenesis of tuberculous meningitis in patients with miliary pulmonary tuberculosis, integrating recent insights. Initially, *Mycobacterium tuberculosis* bacilli spread through the bloodstream from a primary infection site, establishing microscopic foci in the brain’s cortex and meninges, especially in young children. These foci, known as Rich foci, may remain dormant or controlled by the immune system but are more likely to undergo caseation necrosis due to the high bacillary load in miliary tuberculosis. This caseation leads to the discharge of bacilli into the subarachnoid space, initiating tuberculous meningitis.^15^

Secondary dissemination through the bloodstream can also establish caseating foci in the choroid plexus or ventricular walls, contributing to tuberculous meningitis. The immune response to miliary tuberculosis, characterized by widespread inflammation, exacerbates the caseation and discharge processes, promoting the progression to tuberculous meningitis. Paradoxical reactions during antituberculous treatment, due to immune reconstitution inflammatory syndrome, can further expand existing foci or form new lesions. Thus, the high bacillary load and immune dynamics in miliary tuberculosis significantly increase the risk of tuberculous meningitis.^16^

There are certain limitations to our review. Firstly, the exclusion of patients with disseminated tuberculosis, defined as involving two non-contiguous organs, may limit the understanding of central nervous system (CNS) involvement in the broader context of tuberculosis dissemination. This exclusion criterion could lead to an underrepresentation of the spectrum and severity of CNS manifestations in more extensive disease. Additionally, the literature search, while extensive, may still have missed relevant studies, especially those published in less accessible or non-indexed journals. The translation of non-English articles using Google Translate could also introduce translation errors, affecting data accuracy. Finally, the retrospective nature of the included studies limits the ability to draw causal inferences, and the lack of a control group precludes comparison with non-miliary tuberculosis patients.

In conclusion, our review noted various central nervous system involvements in miliary pulmonary tuberculosis, such as tuberculous meningitis and spinal cord lesions. It also highlights common paradoxical manifestations and significant patient improvements following treatment.

## Supporting information

Supplementary item-1

Supplementary item-2

Supplementary item-3

## Declarations

### Conflict of Interest

All authors have no conflict of interest to report.

### The ethical statement

No human or animal subjects were involved so ethical clearance was not taken.

### Funding Declaration

None

## Data Availability

All data lies in the manuscript or supplementary items.

## Notes

### Competing Interest Statement

The authors have declared no competing interest.

### Funding Statement

This study did not receive any funding.

### Summary of Updates

There are no major revisions.

