## Supplementary item-1 for "Central Nervous System Complications in Miliary Pulmonary Tuberculosis: A Systematic Review Of Published Case Reports And Case Series"

**Supplementary item 1: Clinical Characteristics of CNS Manifestations in Miliary Pulmonary Tuberculosis Patients (n=161)**

| **Reference** | **Country** | **Age/sex** | **Co-morbid condition** | **Duration of illness** | **Presenting clinical features** | **Imaging Findings** | **Paradoxical reaction** | **Choroid tuberculoma** | **CSF** | **Microbiological confirmation** | **Drug resistance profile** | **Tissue biopsy/autopsy** | **CNS Manifestations Reported** | **Treatment** | **Outcome** |
| --- | --- | --- | --- | --- | --- | --- | --- | --- | --- | --- | --- | --- | --- | --- | --- |
| Mnaili and Bourazza 2024 | Morocco | 45/F | HIV infection | 1 week | Fever  Respiratory symptoms | Infarct in territory of  the left anterior cerebral artery | NA | NA | Cells=1150 per cumm  Protein= 351 mg/dl  Glucose = 25 mg/dl | Sputum and CSF GeneXpert MTB/RIF = *Mycobacterium*  *tuberculosis* | NA | NA | Tuberculous meningitis | ATT and corticosteroids | Improved |
| Jayadhi Widyakusum et al 2024 | Indonesia | 15/M | NA | 2 days | Altered sensorium  Fever | Leptomeningeal enhancement | NA | NA | Xanthochromia  Cells=111 per cumm  Protein= 1766 mg/dl  Glucose = 46 mg/dl | GeneXpert in gastric fluid= *Mycobacterium*  *tuberculosis* | NA | NA | Tuberculous meningitis | ATT and corticosteroids | NA |
| Kamali et al 2024 | Iran | 12/F | NA | 2 months | Seizures  Focal deficit  Encephalopathy | Multiple tuberculoma of the brain | NA | NA | Cells=50 per cumm  Protein= 370 mg/dl  Glucose = 21 mg/dl | The gastric aspirate culture PCR showed *Mycobacterium tuberculosis* | NA | NA | Tuberculous meningitis with miliary tuberculoma brain | ATT and corticosteroids | Improved |
| Gama e al 2024 | Portugal | 34/F  Immigrant | NA | Sudden | Fever  Left hemiparesis | Multiple tuberculoma of the brain | NA | NA | NA | PCR blood= *Mycobacterium tuberculosis* | NA | NA | Tuberculoma brain | ATT and corticosteroids | Improved |
| El Aggari *et al 2024* | Morocco | 6/F | NA | 2 days | Left hemiparesis | Miliary tuberculoma brain and hydrocephalus | NA | NA | Increased cells only= Cells=25 per cumm | PCR CSF= *Mycobacterium tuberculosis* | NA | NA | Miliary tuberculoma brain | ATT and corticosteroids | Improved |
| Ashizawa et al 2024 | Japan | 52/F | SLE, immunosuppressive therapy | 2 months | Fever  Right  hemiparesis | Tuberculoma of the brain | NA | NA | Cells=93 per cumm  Protein= NA  Glucose = 39 mg/dl  ADA= raised | PCR CSF= *Mycobacterium tuberculosis* | NA | NA | Tuberculous meningitis with tuberculoma brain | ATT and corticosteroids | Improved |
| Tong et al 2023 | China | 14/F | NA | 5 days | Fever  Chest pain  Headache  Vision loss | Miliary tuberculoma brain | Paradoxical optochiasmatic arachnoiditis leading to vision loss  Linezolid neuropathy | NA | Cells= increased  Protein= 133 mg/dl  Glucose = 2.56 mmol/L | CSF GeneXpert MTB/RIF = *Mycobacterium*  *tuberculosis* | Phenotypic MDR | NA | MDR tuberculous meningitis with miliary brain tuberculoma | Linezolid containing MDR ATT | Improved  CSF normal after 6 months |
| Tavares-Júnior et al 2023 | Brazil | 32/F | SLE, immunosuppressive therapy | NA | Fever  Diplopia  Ophthalmoplegia | Thalamic tuberculoma | NA | NA | Cells= 49/cumm  Protein= 95 mg/dl  Glucose = 30 mg/dl | CSF GeneXpert MTB/RIF = *Mycobacterium*  *tuberculosis* | NA | NA | Parinaud syndrome  Tuberculous meningitis with tuberculoma brain | ATT and corticosteroids | Improved |
| Subedi et al 2023 | Nepal | 25/F | NA | 5 months | Fever and cough  Altered sensorium  Seizures | Miliary tuberculoma brain | NA | NA | Lymphocytic pleocytosis  Protein= raised | Sputum  GeneXpert MTB/RIF = *Mycobacterium*  *tuberculosis* | NA | NA | Tuberculous meningitis with miliary tuberculoma brain | ATT and corticosteroids | Improved |
| Samad et al 2023 | France | 42/M  Indian immigrant | NA | 1 month | Fever and respiratory distress  Altered sensorium  Hemiparesis | Miliary tuberculoma brain  Hydrocephalus | NA | NA | Lymphocytic pleocytosis  Protein= raised | BAL PCR= *Mycobacterium*  *tuberculosis* | NA | Liver biopsy= granuloma | Miliary tuberculoma brain | ATT and corticosteroids  ICU  Anti-tumor necrosis  factor agent, infliximab | Improved |
| Muraoka et al 2023 | Japan | 67/F | NA | Sudden | Hemiplegia and dysarthria | Right cerebral infarction and internal carotid artery  Stenosis  Multiple lesions in right hemisphere only | NA | NA | Increased cells and protein | Right-sided endarterectomy= *Mycobacterium tuberculosis* was demonstrated in carotid plague | NA | Right-sided endarterectomy | Tuberculous meningitis | ATT and corticosteroids | Improved |
| Mukherjee et al 2023 | Indian | 21/F | NA | NA | Progressive parkinsonism | Right cerebral infarction with meningeal enhancement | NA | NA | Cells= 82/cumm  Protein= 160 mg/dl  Glucose = 30 mg/dl | CSF GeneXpert MTB/RIF = *Mycobacterium*  *tuberculosis* | NA | NA | Tuberculous meningitis | ATT and corticosteroids | Improved |
| Labbad and Hamzeh 2023 | Syria | 33/F | NA | 5 days | Fever  Headache  Coma | Cerebral venous sinus thrombosis | NA | NA | Cells= 1007/cumm  Protein= 84 mg/dl  Glucose = 29 mg/dl | CSF and Sputum  Culture = *Mycobacterium*  *tuberculosis* | NA | NA | Tuberculous meningitis | ATT and corticosteroids | Improved |
| Idrissa et al 2023 | Guinea | 42/M | NA | 14 days | Fever  Headache  Hemiparesis | Multiple tuberculoma  Intraventricular hemorrhage | NA | NA | Cells= 210/cumm  Protein= 360 mg/dl  Glucose = 25 mg/dl | GeneXpert MTB/RIF CSF = *Mycobacterium*  *tuberculosis* | NA | NA | Tuberculous meningitis with intraventricular hemorrhage | ATT and corticosteroids | Improved |
| Ahmed et al 2023 | Qatar | 35/F | Negative | 1 month | Fever  Headache | Spinal tuberculosis with paravertebral abscess at T8 to L2  Multiple tuberculoma brain | NA | NA | NA | Sputum AFB= positive  Pus from paraspinal abscess AFB= positive | NA | NA | CNS tuberculosis with spinal tuberculosis | ATT and corticosteroids | Improved |
| Wautlet et al 2022 | USA | 51/M | NA | 2 months | Fever and  Headache | Multiple tuberculoma brain | NA | NA | Glucose= 28mg/dl  Proteins= 259 mg/dL  Cells= 143 cells/ml | Sputum and BAL culture = *Mycobacterium*  *tuberculosis* | NA | NA | Tuberculous meningitis with tuberculoma | ATT and corticosteroids | Improved |
| Ohgiya et al 2022 | Japan | 73/M | NA | 1 month | Left hemiparesis  Loss on consciousness | Multiple tuberculoma brain | Brain lesions paradoxically developed while on treatment for miliary tuberculosis | NA | NA | NA | NA | NA | Tuberculous meningitis with tuberculoma | ATT and corticosteroids | NA |
| Letchuman et al 2022 | Kansas | 22/M | NA | Sudden | Seizures  Known case of miliary tuberculosis took ATT for nine months | Multiple tuberculoma brain in temporal region  Paradoxical increase in size of lesion after two months  Vertebral destruction at C5  Tuberculoma in spleen and liver as well | Known case of miliary tuberculosis took ATT for nine months | NA | NA | NA | NA | Liver biopsy = caseating granuloma  Brain biopsy= tuberculous granuloma | Tuberculous meningitis with miliary tuberculoma | Surgical removal of mass lesion  ATT and corticosteroids  Antiepileptic | Improved |
| Khan et al 2022 | UK | 59/M  Immigrant | NA | NA | Seizures  Recurrent adrenal crisis | Multiple tuberculoma brain | Brain lesions paradoxically developed while on treatment for miliary tuberculosis | NA | NA | NA | NA | NA | Possible Tuberculous meningitis with tuberculoma | ATT  and corticosteroids  Antiepileptic | Improved |
| Hirayama et al 2022 | Japan | 70/F | NA | 7 days | Fever  Altered consciousness | NA | Cervical tuberculous lymphadenitis | NA | Glucose= 28 mg/dl  Proteins= 201 mg/dL  Cells= 59cells/ml | CSF  Culture = *Mycobacterium*  *tuberculosis* | NA | Cervical lymph nodes biopsy= *Mycobacterium*  *tuberculosis* | Tuberculous meningitis | ATT and corticosteroids | Improved |
| Christian and Johnston 2022 | USA | 19/M | Crohn’s disease  Adalimumab therapy | 5 days | Fever | Multiple tuberculoma brain | lesions paradoxically there was increase in number and size of lesions and perilesional edema | Multiple choroid tubercles | Normal | Sputum smear= *Mycobacterium*  *bovis* | NA | NA | Tuberculoma brain | ATT and corticosteroids | Improved |
| Ashraf Talesh et al 2022 | Iran | 60/M | COVID-19 | 5 days | Fever  Headache | Miliary brain lesions | NA | NA | Glucose= 25 mg/dl  Proteins= 132 mg/dL  Cells= 01 cells/ml | CSF  Culture = *Mycobacterium*  *tuberculosis* | NA | NA | Tuberculous meningitis | ATT and corticosteroids | Improved |
| de Andrade et al 2022 | Brazil | 3 months/M | NA | NA | Altered consciousness  Bilateral sixth nerve palsy | Multiple intracranial tuberculoma | NA | NA | NA | NA | NA | NA | Intracranial tuberculoma | ATT and corticosteroids | Improved |
| Wong et al 2021 | China | 26/F | Hepatitis C | 2 months | Fever  Cough  Ascites  Paradoxical right hemianopia | Multiple tuberculoma brain | Brain lesions paradoxically developed while on treatment for miliary tuberculosis | NA | NA | Sputum GeneXpert MTB/RIF = *Mycobacterium*  *tuberculosis* | NA | NA | Tuberculoma brain | ATT and corticosteroids | Improved |
| Soni et al 2021  Preprint | India | 28/F | NA | 10 days | Fever  Headache | Miliary brain lesions | NA | NA | Normal | Sputum smear = *Mycobacterium*  *tuberculosis* | NA | NA | Tuberculoma brain | ATT and corticosteroids | Improved |
| Sekkat et al 2021 | morocco | 47/F | Behcet’s disease, on anti-TNF alfa treatment | NA | ATT for four months  Jaundice  Weakness of upper limbs | Miliary brain lesions | Brain lesions paradoxically developed while on treatment for miliary tuberculosis | NA | NA | NA | NA | NA | Tuberculoma brain | ATT and corticosteroids | Improved |
| Sachan et al 2021 | India | 26/M | NA | 1 month | Fever  Vision loss right side  Lymph adenopathy | Spinal tuberculosis in thoracic region | NA | Large exudative retinal detachment with choroidal  granuloma | NA | NA | NA | NA | Spinal tuberculosis | ATT and corticosteroids | Improved |
| Reddy et al 2021 | India | 11/F | NA | 15 days | Fever  Encephalopathy | Multiple tuberculoma brain | NA | Bilateral choroidal  tubercles | NA | Sputum smear= *Mycobacterium*  *tuberculosis* | NA | NA | Tuberculoma brain | ATT and corticosteroids | Improved |
| Pinzon et al 2021 | Indonesia | 39/M | HIV  CD4 count was 84 cells/μL | 2 weeks | Headache  Seizures | Single tuberculoma brain | NA | NA | NA | Sputum smear= *Mycobacterium*  *tuberculosis* | NA | NA | Tuberculoma brain | ATT and corticosteroids | Improved |
| Musa et al 2021 | USA | 21/F  Pregnant | COVID-19 | NA | Took ATT for 2 years  Headache  Seizures  Progressive encephalopathy | Thalamic infarcts  Multiple tuberculoma brain  Basal exudates | Brain lesions paradoxically developed while on treatment | NA | NA | NA | NA | NA | Tuberculous meningitis and miliary tuberculoma brain | ATT and corticosteroids | Improved |
| Kalita et al 2021 | India | 31/M | Prolonged corticosteroids | 2 weeks | Fever  Headache  Seizures  Progressive encephalopathy | Miliary brain lesions | NA | Multiple choroid tubercles | Glucose= 28 mg/dl  Proteins= 152 mg/dL  Cells= 70 cells/ml | CSF  GeneXpert MTB/RIF = *Mycobacterium*  *tuberculosis* | NA | NA | Tuberculous meningitis and miliary tuberculoma brain | ATT and corticosteroids | Improved |
| Duc et al 2021 | Vietnam | 3 months/M | NA | 1 month | Seizures and respiratory insufficiency | Miliary brain lesions | NA | NA | Glucose= 10 mg/dl  Proteins= 300 mg/dL  Cells= 40 cells/ml | PCR CSF was positive for *Mycobacterium tuberculosis* | NA | NA | Tuberculous meningitis and miliary tuberculoma brain | ATT | Died |
| Dormegnie et al 2021 | France | 31/F  Immigrant | On treatment with  Tocilizumab for rheumatoid arthritis | 3 months | NA | NA | NA | Choroidal  tuberculoma | NA | BAL culture= *Mycobacterium tuberculosis* | NA | NA | Choroidal  tuberculoma | ATT and corticosteroids | Improved |
| Cheng et al 2021 | China | 31/F | Following in vitro fertilization | NA  Soon after pre-term delivery | Fever  Headache | Miliary brain lesions | NA | NA | NA | NA | NA | NA | Tuberculous meningitis and miliary tuberculoma brain | ATT | Improved |
| Bouchentouf  2021 | *Morocco* | 31/M | NA | 1 week | Fever  Headache  Hemiparesis | Infarct in left internal capsule | Infarct paradoxically developed 4 days after ATT | NA | Glucose= 23 mg/dl  Proteins= 311 mg/dL  Cells= 150 cells/ml | Sputum and CSF  GeneXpert MTB/RIF = *Mycobacterium*  *tuberculosis* | NA | NA | Tuberculous meningitis | ATT and corticosteroids | Improved |
| Benchekroun et al 2021 | *Morocco* | 32/M | NA | 2 weeks | Vision loss right side | Miliary tuberculoma  Optic nerve tuberculoma | Optic nerve tuberculoma paradoxically developed after ATT for abdominal tuberculosis | NA | NA | NA | NA | NA | Miliary tuberculoma | ATT and corticosteroids | Died due to liver failure |
| Bǎiceanu et al 2021 | Romania | 42/M | Alcoholic hepatic cirrhosis | Sudden | Fever  Loss of consciousness  Hemiparesis | Miliary brain lesions | Paradoxical increase in number of lesions | NA | Glucose= 70 mg/dl  Proteins= 121 mg/dL  Cells= < 5 cells/ml | BAL GeneXpert MTB/RIF= *Mycobacterium*  *tuberculosis* | NA | NA | Miliary tuberculoma | ATT and corticosteroids | Improved |
| Greenberg e et al 2021 | USA | 11/F  Immigrated | NA | NA | Fever  Altered consciousness | Thalamic infarcts  Spinal tuberculosis extending T10 and T11 vertebral bodies | NA | NA | Glucose = 20 mg/dL  Proteins= 92  mg/dL  Cells=945 /mm3 | Sputum and gastric aspirates smear and culture= *Mycobacterium* | NA | NA | Tuberculous meningitis | ATT and corticosteroids | Improved |
| Velásquez-Rimachi et al 2020 | Peru | 21/F | NA | 3 months | Headache  Ataxia  Vision loss  Altered sensorium | Multiple tuberculoma brain  Multiple spinal cord tuberculoma | NA | NA | Proteins= 84 mg/dL  Cells= < 2 cells/ml | Negative | NA | NA | Tuberculous meningitis and miliary tuberculoma brain and spinal cord | ATT and corticosteroids | Improved |
| Vasconcelos et al 2020 | Portugal | 47/F | NA | 1 day | Altered sensorium | Miliary brain lesions | NA | NA | Glucose= 37 mg/dl  Proteins= 128 mg/dL  Cells= < 120 cells/ml | BAL = *Mycobacterium*  *tuberculosis* | NA | NA | Tuberculous meningitis and miliary tuberculoma brain | ATT and corticosteroids | Improved |
| Tetsuka et al 2020 | Japan | 91/F | NA | 1 day | Altered sensorium | Miliary brain lesions | NA | NA | Glucose= 12 mg/dl  Proteins= 374 mg/dL  Cells= < 180 cells/ml | Sputum and gastric fluid smear= *Mycobacterium*  *tuberculosis* | NA | NA | Tuberculous meningitis and miliary tuberculoma brain | ATT and corticosteroids | Died |
| Sivakami et al 2020 | India | 12/F | NA | 15 days | Fever  Headache | Multiple tuberculoma brain | NA | Choroid tubercles | NA | NA | NA | NA | Miliary tuberculoma brain | ATT | Improved |
| Rahman et al 2020 | India | 17/M | NA | 3 months | Fever  Headache | Miliary brain lesions  Lesion in liver | NA | NA | Glucose= 24 mg/dl  Proteins= 123 mg/dL  Cells= < 100 cells/ml | CSF  GeneXpert MTB/RIF = *Mycobacterium*  *tuberculosis* | NA | NA | Tuberculous meningitis and miliary tuberculoma brain | ATT and corticosteroids | Improved |
| Meriem et al 2020 | Tunisia | 35/F  Pregnant | NA | 1 month | Fever  Headache | Meningeal enhancement | NA | NA | NA | NA | NA | NA | Possible CNS tuberculosis | ATT and corticosteroids | Improved |
| Ish et al 2020 | India | 16/F | NA | 2 weeks | Vision loss left | NA | NA | Choroidal  tuberculoma | NA | Sputum  GeneXpert MTB/RIF = *Mycobacterium*  *tuberculosis* | NA | NA | Choroidal tuberculoma | NA | NA |
| Esposito et al 2020 | USA | 30/M | Negative | NA | Headache | Multiple tuberculoma brain  Paravertebral abscess  Pott’s spine | NA | NA | Glucose = 14 mg/dl  Proteins= 351 mg/dL  Cells= < 91 cells/ml | CSF/PUS culture = *Mycobacterium*  *tuberculosis* | NA | NA | Tuberculous meningitis and spinal tuberculosis | ATT and corticosteroids | Improved |
| St Cyr and Starke 2020 | USA | 3 months/M | Negative | 1 month | Fever and cough | Miliary brain lesions | NA | NA | Glucose= 53 mg/dl  Proteins= 36 mg/dL  Cells= 5 cells/ml | BAL revealed *Mycobacterium tuberculosis* | NA | NA | Miliary tuberculoma brain | ATT | Improved |
|  |  | 8  months/M | Negative | Several weeks | Fever and cough | Miliary brain lesions | NA | NA | Normal | Sputum revealed *Mycobacterium tuberculosis* | NA | NA | Miliary tuberculoma brain | ATT | Improved |
| Sánchez-Códez et al 2019 | Germany | Infant/F | NA | 7 months | Fever  Seizures | Miliary brain lesions  Basal ganglionic infarct | NA | NA | Lymphocytic pleocytosis | CSF  GeneXpert MTB/RIF = *Mycobacterium*  *tuberculosis* | NA | NA | Tuberculous meningitis  Miliary tuberculoma brain | ATT | Improved |
| Hansen et al 2019 | Denmark | 54/M | NA | NA | Headache | Miliary brain lesions | NA | NA | Normal | BAL revealed *Mycobacterium tuberculosis* | NA | NA | Miliary tuberculoma brain | ATT and corticosteroids | Improved |
| Boubnan et al 2019 | Morocco | 36/F | NA | 3 days | Right vision loss | Multiple brain tuberculoma | NA | A large choroidal  tuberculoma | NA | NA | NA | NA | Brain tuberculoma and choroidal tuberculoma | ATT | Affected eye was removed |
| Amin et al 2019 | USA | 32/F  Immigrant | In vitro fertilization | 1 week | Headache | Multiple brain tuberculoma | NA | NA | Normal | NA | NA | Endometrial and liver biopsy= caseating granulo,ma AFB was demonstrated | Brain tuberculoma and choroidal tuberculoma | ATT | Improved |
| Zhan et al 2018 | China | 39/F | Negative | 2 months | Fever  Headache | Miliary brain tuberculoma  Tuberculous liver abscess | Brain lesions paradoxically developed while on ATT of two months | NA | Glucose= 3.0 mmol/L  Proteins= 28 mg/dL  Cells= 80 cells/ml | NA | Streptomycin resistant | Lung biopsy = tuberculous granuloma that demonstrated *Mycobacterium tuberculosis* | Miliary brain tuberculoma | ATT | Improved |
| Vélez-Tirado and Cote-Orozco 2018 | Colombia | 7/M | NA | 15 days | Headache  Altered sensorium  Seizures  Papilledema  Hemiparesis | Frontal lobe tuberculoma | NA | NA | NA | Bain biopsy specimen PCR – Positive, culture negative | NA | Brain biopsy= caseating granuloma | Tuberculoma brain | ATT and corticosteroids | Improved |
| Sharma et al 2018 | India | 5/M | NA | 2 months | Fever | Multiple tuberculoma brain  Multiple spinal cord tuberculoma  Meningeal enhancement | NA | Choroidal tubercles | Glucose= 26 mg/dl  Proteins= 48 mg/dL  Cells= 140 cells/ml | NA | NA | NA | Tuberculous meningitis  Miliary tuberculoma brain | ATT | Improved |
| Saraswat et al, 2018 | India | 5/ NA | NA | 20 days | Fever, headache, abnormal movement of right eye | Multiple tuberculoma, intraventricular tuberculoma | NA | NA | NA | NA | NA | NA | Tuberculoma | ATT | NA |
| Parija et al, 2018 | India | 28/M | NA | 2 days | Headache, tingling sensation of right upper limb, drooping of left eyelid, fever | Midbrain tuberculoma | NA | NA | NA | NA | NA | NA | Midbrain tuberculoma – Weber syndrome | ATT and corticosteroids | Improved |
| Rodriguez et al, 2018 | Peru | 63/F | Diabetes mellitus and chronic kidney disease | 3 weeks | Headache, drowsiness, hypoactivity, anorexia, asthenia | Miliary tuberculoma | NA | NA | Turbid CSF, Protein = 272.3 mg/dl  Glucose = 18  TLC = 310 (85% polymorphonuclear)  ADA = 38.2 U/L | NA | NA | NA | Tuberculous meningitis with CNS tuberculosis | ATT and corticosteroids | Died |
| Macauley et al, 2018 | USA | 40/F | Lupus Nephritis on steroids and mycophenolate mofetil | 4 days | Fever, sinus pressure, chills, rigors and occipital headache | Leptomeningeal enhancement | NA | NA | Protein = 137 mg/dl  Glucose < 10 mg/dl  WBC = 426 (50% polymorphs, 46% lymphocytes, 5% monocytes) | CSF AFB positive  CSF culture positive for *Mycobacterium tuberculosis* | NA | NA | Tuberculous meningitis with miliary pulmonary tuberculosis | NA | Improving |
| Ko et al, 2018 | Korea | 72/F | NA | 2 Months | Generalized weakness, poor oral intake, dysarthria, sluggish speech | Multiple tuberculoma | New lesions on brain MRI 4 weeks after starting ATT | NA | Protein = 32 mg/dl  Glucose = 81 mg/dl  TLC = Nil | CSF and bronchoalveolar lavage PCR positive for *Mycobacterium Tuberculosis* | Drug sensitive to primary line anti-tubercular drugs | Right renal biopsy showed granulomatous inflammation and TB PCR positive for *Mycobacterium Tuberculosis* | Miliary tuberculosis, multiple intracranial tuberculoma with a paradoxical response to ATT | ATT and corticosteroids | Improved |
| Kim et al, 2018 | Korea | 65/F | Post-Kidney transplant | 2 Weeks | Cough | Intracranial and spinal tuberculoma | Paradoxical reaction in form of progressive paraparesis 2 weeks after starting ATT for miliary pulmonary tuberculosis | NA | NA | Bronchioalveolar lavage PCR positive for *Mycobacterium Tuberculosis* | Drug sensitive to primary line anti-tubercular drugs | Surgical resection of spinal lesion showed granulomatous inflammation and positive AFB | Drug susceptible miliary tuberculosis with spinal and cerebral tuberculoma with paradoxical response to ATT | ATT with corticosteroids | Improved |
| Balal et al, 2018 | Turkey | 32/F | NA | 15 days | Numbness in right hand and right side of face | Multiple intracranial tuberculoma | NA | NA | Protein = 43 mg/dl  Glucose = 53 mg/dl  TLC = Nil | CSF PCR/Culture negative for *Mycobacterium Tuberculosis*  QuantiFERON test positive | NA | NA | Miliary tuberculosis with CNS involvement from salpingitis due to immunosuppression caused by pregnancy | NA | NA |
| Pal et al, 2017 | India | 15/M | NA | 2 Months | Cough, low-grade fever | NA | NA | Choroid tubercles present | NA | NA | NA | NA | Miliary tuberculosis with choroid tubercles in an immunocompetent host. | ATT with corticosteroids | Improved |
| Soria et al, 2017 | Argentina | 24/M | Severe malnutrition | 2 Months | Weight loss, asthenia and pre-cervical mass | Brain abscess frontal lobe, cervical and lumbar abscess | NA | Choroidal lesions with Subretinal tubercular abscess | NA | NA | NA | AFB positive in pre-cervical mass FNAC | Miliary pulmonary tuberculosis, frontal lobe abscess, cervical and lumbar spinal abscess, hepatic abscess, subretinal tubercular abscess | ATT | Improved |
| Madhyastha et al, 2017 | India | 20/F | NA | NA | One episode of generalized tonic-clonic seizure | Multiple ring-enhancing lesions | NA | NA | Protein = 84 mg/dl  Glucose = 43 mg/dl  TLC = Normal cell count | AFB positive in pelvic fluid collection | Sensitive to isoniazid and rifampicin | NA | Miliary tuberculosis, multiple brain tuberculoma, pelvic collection in left lumbar and iliac fossa. | ATT with corticosteroids | Improved |
| Laleona et al, 2017 | Spain | 9/F | Juvenile rheumatoid arthritis on monoclonal antibody | NA | Fever, left inguinal lymphadenitis and left knee arthritis | Brain tuberculomas | NA | NA | NA | Molecular diagnosis of left synovial biopsy, gastric and bronchial aspirate revealed *Mycobacterium tuberculosis* | Pan-susceptible *Mycobacterium tuberculosis* | Synovial biopsy left knee chronic synovitis with granulomatous necrotizing inflammation | Reactivated miliary tuberculosis, cerebral tuberculoma, left knee tubercular arthritis secondary to adalimumab for Juvenile rheumatoid arthritis | ATT with corticosteroids | Improved |
| Garg et al, 2017 | India | 36/F | NA | 2 Months | Fever, headache, weakness of right side | Miliary brain tuberculoma | NA | NA | Protein = 24 mg/dl  Glucose = 59 mg/dl  TLC = 3 cells/cumm | NA | NA | NA | Miliary brain tuberculoma and miliary pulmonary tuberculosis | ATT with corticosteroids | Improved |
| Emamzadehfard et al, 2017 | USA | 28/M | HIV disease | NA | Ataxia | Ring enhancing lesions supratentorial brain with acute infarction of anterior body of corpus callosum | NA | NA | NA | NA | NA | Na | Brain tuberculoma, miliary pulmonary tuberculosis with HIV disease | ATT with highly active anti-retroviral therapy | Improved |
| Zhao et al, 2016 | China | 27/F | NA | 2 Months | Fever, cough, chills, headache, vomiting | Multiple ring enhancing lesions in brain, brain stem and ventricle | Brain tuberculoma diagnosed 2 months after starting ATT for pulmonary miliary tuberculosis | NA | Protein = 0.66 g/l  Glucose = 2.38 mmol.l  TLC = dominated by neutrophil and lymphocytes | CSF GeneXpert positive for *Mycobacterium tuberculosis* | NA | NA | Intracranial miliary tuberculoma with tuberculous meningitis and miliary tuberculosis | ATT with corticosteroids | Improved |
| Viel-Thériault et al, 2016 | Canada | 13/F | NA | NA | Cough, weight loss, night sweats, abdominal pain, severe headache with morning emesis | Multiple brain tuberculomas with pseudo abscesses and meningeal enhancement | New brain tuberculoma after initiating ATT | Suspected two retinal lesions | Protein = 0.83 g/L  Glucose = 1.1 g/L  Moderate pleocytosis 59% lymphocytes | Gastric aspirate and sputum positive for *Mycobacterium tuberculosis*  CSF PCR negative for *Mycobacterium tuberculosis* | Drug sensitive to usual TB drugs | NA | Miliary tuberculosis with brain tuberculoma and disseminated disease with paradoxical response | ATT with corticosteroids and thalidomide | Improved |
| Rali et al, 2016 | USA | 45/M | NA | 2 Weeks | Headache, agitation, cough, seizure | Brain and brainstem tuberculoma | NA | NA | CSF showed lymphocyte predominance | CSF, bronchioalveolar lavage positive for *Mycobacterium tuberculosis* | Susceptible to standard first-line drugs | NA | Tuberculous meningitis with multiple tuberculoma, paradoxical response to ATT and miliary pulmonary tuberculosis | ATT with Corticosteroids (added after paradoxical response) | Improved |
| Parmaksiz et al, 2016 | Turkey | 30/F  Pregnancy | NA | NA | Fever, vaginal bleeding | Tubercular meningoencephalitis | NA | NA | Protein = 6.8g/l  Glucose = 5 mmol/L  TLC = 800 cells/cumm | Lung biopsy tissue positive for  *Mycobacterium tuberculosis* | Resistant to isoniazid, rifampicin, ethambutol and streptomycin | Lung tissue – necrotizing granulomatous inflammation | Multi-drug resistant pulmonary miliary tuberculosis, tuberculous meningitis with disseminated disease | Para-amino salicylic acid, cycloserine, protionamide, moxifloxacin, amikacin and pyrazinamide | Improved |
| Baudel et al, 2015 | France | 22/M | NA | 6 Months | First episode of seizure, asthenia, headache, cough, and limping for 6 months | Multiple brain tuberculoma, Sacroiliitis | NA | NA | Protein = 43g/l  Glucose = 1.2 mmol/l  TLC - 119 per milliliter (67 %  neutrophils, 14 % lymphocytes) | CSF positive for AFB  GeneXpert positive for *Mycobacterium tuberculosis* | Drug sensitive to Rifampicin | NA | Brain tuberculoma with tuberculous meningitis, sacroiliitis, miliary pulmonary tuberculosis | ATT | NA |
| Wang et al, 2015 | USA | 60/M | Chron’s disease on infliximab | 2 Months | Fever, fatigue and weight loss | Multiple brain tuberculoma | NA | NA | Elevated protein, low glucose and negative for mycobacterial culture | Sputum culture, colonic biopsy PCR positive for *Mycobacterium tuberculosis* | Drug sensitive to rifampicin and isoniazid | NA | Colonic biopsy showed granulomas with positive AFB | ATT | Died |
| Tanaka et al, 2015 | Japan | 64/F | Rheumatoid arthritis on adalimumab | NA | Fever, neck swelling, CNS symptoms – headache, nauseas started 3 weeks after ATT | Multiple brain tuberculoma, miliary nodules in lungs, spleen and kidneys | Brain tuberculoma after 3 weeks of starting ATT | NA | Protein = 62 mg/ml  Glucose = 33 mg/dl  TLC - 23 per cumm (68 %  neutrophils, 32 % lymphocytes) | Sputum and urine PCR and cultures positive for *Mycobacterium tuberculosis* | Drug sensitive to all anti-TB drugs | NA | Multiple tuberculoma with tuberculous meningitis with miliary disseminated disease | ATT with corticosteroids | Improved |
| Prasad et al, 2015 | India | 28/M | NA | 1 Month | Progressive weakness of both legs | Multiple brain and intramedullary spinal cord tuberculoma | NA | NA | NA | NA | NA | NA | Multiple brain and spinal cord tuberculoma with miliary pulmonary tuberculosis | ATT with corticosteroids | Improved |
| Islam et al, 2015 | India | 5/F | NA | 10-12 Days | Fever, cough, nausea, vomiting, headache, visual disturbances, photophobia | Multiple granulomatous lesions in right fronto-parietal and left cerebellar region | NA | NA | NA | NA | NA | NA | Brain tuberculoma with miliary pulmonary tuberculosis | NA | NA |
| Hsieh et al, 2015 | Taiwan | 29/F | Noonan syndrome | 2 Months after initiating ATT for miliary pulmonary tuberculosis | Bitemporal headache | Multiple brain abscess with meningitis | Brain tuberculoma appeared after initiating ATT for pulmonary tuberculosis | NA | NA | Stereotactic aspirate from brain lesion positive for AFB and PCR positive for *Mycobacterium tuberculosis* | NA | NA | Tubercular brain abscess with miliary pulmonary tuberculosis | ATT with corticosteroids | Improved |
| Gunbatar et al, 2015 | Turkey | 58/F | End stage renal disease with adult polycystic kidney disease | 3 Days | Fever, weakness, impairment of consciousness | Minimal enlargement of lateral ventricles | NA | NA | Protein = 89 mg/dl  Glucose = 63 mg/dl  Chloride = 128 mmol/L | NA | NA | Left supraclavicular lymph node biopsy revealed caseating granulomatous lymphadenitis | Miliary tuberculosis with tubercular meningitis and tubercular lymphadenitis | ATT | NA |
| Gamell et al, 2015 | Tanzania | 9/M | HIV Disease | NA | Weight loss, fatigue, night sweats – developed left hemiplegia and inguinal lymphadenopathy on continuation phase of ATT | NA (CT Scan not available at hospital) | NA | NA | Protein = low  Glucose = 1.1 mmol/L  TLC = 50 /MicroL | GeneXpert in CSF and lymph node aspirate positive for *Mycobacterium tuberculosis* | CSF sensitive to rifampicin, lymph node aspirate resistant to rifampicin | NA | HIV infection, WHO stage 4, disseminated tuberculosis, rifampicin-susceptible tuberculous meningitis and rifampicin-resistant tuberculous lymphadenitis, miliary pulmonary tuberculosis severe malnutrition | Rifampicin, isoniazid, pyrazinamide, ethambutol, streptomycin, ciprofloxacin, metronidazole, cotrimoxazole and corticosteroids | Died |
| Diguvinti et al, 2015 | India | 32/M | NA | 10 Days | Unsteadiness of gait, swaying, slurred speech, heaviness of both upper/lower limbs | Multiple ring-lesions in cerebral, cerebellar hemispheres and brainstem and spinal cord | NA | NA | Protein = 37 mg/dl  Glucose = 52 mg/dl  TLC = 4 /cu mm | CSF PCR positive for *Mycobacterium tuberculosis* | NA | NA | Multiple cerebral, brain stem and spinal cord tuberculoma with miliary pulmonary tuberculosis | ATT with corticosteroids | Improved |
| Arora et al, 2015 | India | 7/M | NA | 6 Weeks | Fever, frontal headache, vomiting, decreased appetite | Multiple brain tuberculoma | New brain lesions on continuation phase of treatment | Choroid tubercles present | CSF not done | NA | NA | NA | Multiple brain tuberculoma with miliary pulmonary tuberculosis with paradoxical worsening | ATT with corticosteroids | Improved |
| Yoo et al 2014 | Korea | 27/M | HIV Disease  CD4= 55 cells/μL  On ART | 2 weeks | Fever  Rash  Encephalopathy | NA | NA | NA | Cells = 1,258/mm3 | Sputum PCR/ culture= *Mycobacterium tuberculosis*  PCR in CSF= *Mycobacterium tuberculosis* | MDR resistance including resistance to quinolone  Later  resistance to ethambutol  and pyrazinamide | Skin biopsy= *Mycobacterium tuberculosis* | MDR tuberculous meningitis | Modified ATT | Improved |
| Tsai et al 2014 | Taiwan | 20/F | NA | 1 month | Fever  Headache | NA | Multiple tuberculoma paradoxically developed | NA | NA | Sputum culture= *Mycobacterium tuberculosis* | NA | NA | Multiple brain tuberculoma | ATT with corticosteroids | Improved |
| Ranganath et al 2014 | USA | 53/M  Immigrant | NA | 1 week | Fever, cough and headache  Altered sensorium | Basal exudates | NA | NA | Glucose= 34 mg/dl  Proteins= 142 mg/dL  Cells= 313 cells/ml | CSF and sputum culture= *Mycobacterium tuberculosis* | NA | Testicular biopsy= tuberculous granuloma that demonstrated *Mycobacterium tuberculosis* | Tuberculous meningitis | ATT and corticosteroids | Improved |
| Kumar et al 2014 | India | 3 months/M | NA | 2 months | Fever  Seizures | Meningeal enhancement  Hydrocephalus  Ventriculitis | NA | NA | Glucose= 24 mg/dl  Proteins= 119 mg/dL  Cells= 400 cells/ml | NA | NA | Post-mortem biopsy brain = caseating granuloma with *Mycobacterium tuberculosis* | Tuberculous meningitis | ATT | Died |
| Kenzaka and Noda 2014 | Japan | 69/M | NA | 1 month | Fever  Progressive encephalopathy | Multiple tuberculoma | NA | NA | NA | Sputum= *Mycobacterium tuberculosis* | NA | NA | Multiple tuberculoma brain | ATT | Improved |
| Iqbal et al 2014 | India | 24/M | NA | 2 days | Left brachial monoplegia  Headache and fever | Multiple tuberculoma | NA | NA | Glucose= 50 mg/dl  Proteins= 99 mg/dL  Cells= 1 cells/ml | NA | NA | NA | Tuberculous meningitis  Tuberculoma brain | ATT and corticosteroids | Improved |
| Hilal et al 2014 | USA | 29/M  Immigrant | NA | NA | Altered sensorium | Normal | NA | NA | Glucose= 94  mg/dl  Proteins= 85 mg/dL  Cells= 13 cells/ml | CSF  GeneXpert MTB/RIF = *Mycobacterium*  *Tuberculosis*  AFB in urine and sputum | NA | NA | Tuberculous meningitis | ICU  ATT and corticosteroids | Improved |
| Sönmez et al 2013 | Turkey | 54/M | NA | NA | Paraparesis | Multiple tuberculoma brain  Tuberculoma spinal cord | NA | NA | Glucose = 58  mg/dl  Proteins= 66 mg/dL  Cells= 0 cells/ml | CSF  PCR = *Mycobacterium*  *Tuberculosis* | NA | NA | Tuberculoma brain and spinal cord | ATT and corticosteroids | Improved |
| Smith et al 2013 | Australia | 17 months/M  immigrated | NA | 2 months | Fever  Seizures | Miliary brain tuberculoma | NA | NA | Glucose = 27  mg/dl  Proteins= 100 mg/dL  Cells= 29 cells/ml | QuantiFERON-TB Gold test= positive  CSF culture= *Mycobacterium*  *Tuberculosis* | NA | NA | Tuberculous meningitis | ATT and corticosteroids  Ventriculoperitoneal shunt  ICU | Improved |
| Patel et al 2013 | India | 5/M | Malnourished | Sudden | Patient had miliary pulmonary tuberculosis and on ATT  Paradoxically developed seizures | Miliary brain tuberculoma | Paradoxically developed after 3 weeks of ATT | NA | NA | FNAC cervical lymph node= *Mycobacterium*  *Tuberculosis* | NA | Lymph node biopsy= tuberculous granuloma | Miliary tuberculoma brain | ATT and corticosteroids | Improved |
| Pasticci et al 2013 | Italy | 45/M | HIV positive  On ART  CD4= 144/CUMM | 2 days | Right hemiplegia after 5^th^ day of ATT | Left parieto-occipital infarct  MCA occlusion | NA | NA | Glucose = 25  mg/dl  Proteins= 273 mg/dL  Cells= 150  cells/ml | BAL smear= *Mycobacterium*  *Tuberculosis* | NA | NA | Middle cerebral artery occlusion leading to stroke | ATT and corticosteroids  Aspirin | Improved |
| Ishiwada et al 2013 | Japan | 3 month/F | NA | 3 weeks | Respiratory symptoms | Miliary brain tuberculoma | NA | NA | Normal | Gastric aspirates smear= *Mycobacterium*  *Tuberculosis* | Isoniazid-and streptomycin-resistant | NA | Miliary brain tuberculoma | ATT | Improved |
| De La Riva et al 2013 | Spain | 35/F | NA | 1 week | Altered consciousness  Fever  41 days after taking ATT  Blurred vision, diplopia. And vomiting | Miliary brain tuberculoma | Paradoxically increase in size of the lesions along with increase in perilesional edema. | NA | Glucose = 77  mg/dl  Proteins= 58 mg/dL  Cells= 25  cells/ml | Sputum= *Mycobacterium tuberculosis* | NA | NA | Miliary brain tuberculoma | ATT and corticosteroids  Thalidomide | Improved |
| Winklhofer and Kollias  2012 | Switzerland | 49/F | Multiple myeloma  high-dose chemotherapy and bone marrow  transplantation. | NA | Tongue swelling | A tuberculoma in the pons. | NA | NA | Lymphocytic pleocytosis | Sputum PCR= *Mycobacterium tuberculosis* | NA | Tongue biopsy= tuberculous granuloma with AFB | Brain stem tuberculoma | ATT and corticosteroids | Improved |
| Verma et al 2012 | India | 25/M | NA | 6 weeks | Paraparesis  Diplopia  Ptosis | Meningeal enhancement and hydrocephalus  Spinal tuberculosis L1-L4 with paravertebral abscess | NA | NA | Glucose = 58  mg/dl  Proteins= 180 mg/dL  Cells= 150  cells/ml | Sputum smear= *Mycobacterium tuberculosis* | NA | CT-guided biopsy of the vertebral lesion= tuberculous granuloma with AFB | Tuberculous meningitis with spinal tuberculosis | ATT and corticosteroids | Improved |
| Sundaram et al 2012 | UK | 37/F | HIV-positive  On ART  CD4 count= 10 cells/mm3 | 6 weeks | Fever  Headache | Epidural abscess  causing irregular cord compression between C7 and T10/T11 | 6 years later  Multiple brain tuberculoma  A multiloculated syrinx from  the T2 to T11 levels | NA | NA | BAL and CSF culture= *Mycobacterium tuberculosis* | Resistant to both isoniazid  and streptomycin | Epidural biopsy PCR= *Mycobacterium tuberculosis*  CT-guided abdominal lymph node biopsy= tuberculous granuloma | Tuberculous meningitis with spinal tuberculosis | ATT and corticosteroids  Neurorehabilitation | Improved |
| Şen et al 2012 | Turkey | 45/M | NA | NA | Headache  Left vision loss | Multiple brain tuberculoma | NA | A choroidal tubercle | NA | BAL smear= *Mycobacterium tuberculosis* | NA | NA | Multiple brain tuberculoma with choroidal tuberculoma | NA | NA |
| Morioka et al 2012 | Japan | 78/F | NA | 1 week | Fever  Hemiparesis left  Altered consciousness | Multiple small infarcts | After 5 weeks of ATT, she had another infarct | NA | Glucose = 53  mg/dl  Proteins= 92 mg/dL  Cells= 18  cells/ml | NA | NA | NA | Tuberculous meningitis | ATT and corticosteroids | Improved |
| Jorge et al 2012 | Mexico | 20/M | Rheumatoid Arthritis on Long term steroids and immunosuppressive  Infliximab  HLAB27 positive | NA | Fever  Headache  After 2 months of treatment | Multiple brain tuberculoma | 2 months later  vasculitis at both middle cerebral arteries | NA | Glucose = 9  mg/dl  Proteins= 293 mg/dL  Cells= 405  cells/ml | CSF and urine cultures= *Mycobacterium tuberculosis* | NA | NA | Tuberculous meningitis  Multiple brain tuberculoma | ATT and corticosteroids  VP shunt  Infliximab | Improved at At 39 months of follow-up |
| Gönen et al 2012 | Turkey | 22/M | Behcet’s disease  on Long term steroids and immunosuppressive | 10 days | Fever  Headache  Epididymitis | Miliary brain tuberculoma  Spinal tuberculosis in lumbar vertebra | NA | NA | Glucose = 27  mg/dl  Proteins= 200 mg/dL  Cells= 20  cells/ml | Scrotal FANC= *Mycobacterium tuberculosis* | NA | NA | Tuberculous meningitis  Multiple brain tuberculoma  Spinal tuberculosis | ATT and corticosteroids | Improved |
| Das et al 2012  (A report of 5 cases, 2 had miliary tuberculosis) | India | 22/F | NA | 2 months | Fever  Headache | Miliary brain tuberculoma | Miliary brain tuberculoma  paradoxically developed | NA | Normal | NA | NA | NA | Miliary brain tuberculoma | ATT and corticosteroids | Improved |
|  |  | 10/F | NA | 3 months | Fever  Headache  Back pain  On ATT for 2 months, she had seizures | Spinal tuberculosis | A large tuberculoma paradoxically developed | NA | Lymphocytic pleocytosis | NA | NA | NA | Tuberculous meningitis  A large tuberculoma | ATT and corticosteroids | Improved |
| Chaudhry et al 2012 | Pakistan | 31/F | NA | 1 month | Fever  After 22 days of ATT she developed paraplegia | Miliary brain tuberculoma | Miliary brain tuberculoma  paradoxically developed  in brain and brainstem | Bilateral choroid eye lesions | Lymphocytic pleocytosis | Sputum smear= *Mycobacterium tuberculosis* | NA | NA | Tuberculous meningitis  A large tuberculoma | ATT and corticosteroids | Improved |
| Yasar et al 2011 | Turkey | 14/F  Immigrant | NA | 10 days | Fever  Headache  Altered sensorium  Seizures | Multiple brain tuberculoma | NA | NA | Glucose = 13  mg/dl  Proteins= 243 mg/dL  Cells= 57  cells/ml | CSF  GeneXpert MTB/RIF and culture = *Mycobacterium*  *Tuberculosis* | NA | NA | Tuberculous meningitis  Multiple brain tuberculoma | ATT and corticosteroids  ICU | Improved with persistent motor deficits |
| Yasar et al 2011 | Turkey | 23/F | Negative | 15 days | Fever  Headache  Seizures | Multiple brain tuberculoma | Paradoxical enlargement of brain lesions were noted | Bilateral choroid  tubercles | NA | NA | NA | NA | Tuberculous meningitis  Multiple brain tuberculoma | ATT and corticosteroids  ICU | Improved |
| Undrakonda and Umakanth 2011 | India | 20/M | NA | NA | Fever  Diplopia | Pontine tuberculoma | NA | NA | NA | Sputum smear= *Mycobacterium tuberculosis* | NA | NA |  | ATT and corticosteroids | Died on 7thday |
| Papastefanou and Cohen 2011 | UK | 24/M  Immigrant | NA | NA | Vision loss | Multiple brain tuberculoma | NA | Choroidal mass with retinal  detachment | NA | NA | NA | NA | A large choroidal tuberculoma | ATT | Improved |
| Padhi et al 2011 | India | 24/F | NA | 4 months | Vision loss right  Fever  Headache | Tuberculoma of optic disc  Multiple brain tuberculoma | NA | Retinal  detachment | NA | NA | NA | NA | Tuberculoma of optic disc | ATT and corticosteroids | Improved |
| Ikegame et al 2011 | Japan | 63/M | Chronic myeloid  Leukemia  On immunosuppressive therapy | 1 month | Fever  Had miliary tuberculosis  ATT started  After 2 months  Headache and vomiting | Normal | Paradoxical tuberculous meningitis | NA | Lymphocytic pleocytosis | Sputum smear= *Mycobacterium tuberculosis*  Positive PCR results for Pneumocystis  jiroveci | INH-resistant | NA | Tuberculous meningitis | ATT | Improved |
| Hess et al 2011 | Germany | 17/M | Refractory synovitis,  acne, pustulosis, hyperostosis, and osteitis (SAPHO) syndrome  On anti-TNF treatment adalimumab | 5 days | Ataxia  Dysarthria  Fever  Altered sensorium  Dysnoea  Septic shock | Multifocal osteomyelitis  Miliary brain tuberculoma | Paradoxical tuberculous meningitis | NA | Glucose = 05  mg/dl  Proteins= 396 mg/dL  Cells= 275  cells/ml | Interferon-γ-release assay (ELISpot) for tuberculosis= positive  BAL PCR= *Mycobacterium tuberculosis*  CSF culture= *Mycobacterium tuberculosis* | NA | NA | Tuberculous meningitis  Miliary brain tuberculoma | ATT and corticosteroids  Thalidomide  ICU | Improved |
| Seif et al 2010 | USA | 50/F | NA | NA | Fever  Altered consciousness  Left otorrhea | Multiple tuberculoma with ventriculitis  Vertebral lesions (Pott’s spine)  Tuberculoma in liver and spleen | Paradoxical optochiasmatic arachnoiditis | NA | NA | Aspirate from vertebral lesion= *Mycobacterium tuberculosis* | NA | NA | Tuberculous meningitis  And brain tuberculoma | ATT and corticosteroids | Not improved |
| Rey and Sousa 2010 | Brazil | 4/F | Negative | 2 months | Fever | A large abscess in posterior fossa | NA | NA | NA | Aspirate from lymph node= *Mycobacterium tuberculosis* | NA | NA | Tuberculous abscess | ATT | Improved |
| Ottaviani et al 2010 | France | 39/M | Infliximab  Anti-TNF-a therapy | NA | Fever  Progressive radicular pain | Spinal intramedullary tuberculoma | NA | NA | Normal | NA | NA | NA | Spinal intramedullary tuberculoma | ATT and corticosteroids | Improved |
| Krygowski et al 2010 | USA | 2/M | NA | 3 week | Fever  Choreoathetosis and unsteady gait | Multiple brain tuberculoma | NA | NA | NA | Gastric aspirate= *Mycobacterium tuberculosis* | NA | Brain biopsy= tuberculous granuloma | Multiple brain tuberculoma | ATT | Improve |
| Aslan et al 2010 | Turkey | 60/F | NA | 1 month | Fever  cervical lymphadenopathy  Spinal tuberculosis  Headache  Altered sensorium | Multiple brain tuberculoma | NA | NA | Glucose = 42  mg/dl  Proteins= 200 mg/dL  Cells= 40  cells/ml | CSF culture= *Mycobacterium tuberculosis* | NA | NA | Tuberculous meningitis  brain tuberculoma | ATT and corticosteroids | Improved |
| Vishnubhotla et al 2009 | India | 55/M | Post-transplant immunosuppression | 4 months | Fever  Headache  Altered sensorium | Multiple brain tuberculoma  And one in right orbit | NA | NA | NA | Urine= *Mycobacterium tuberculosis* | NA | NA | Brain tuberculoma | ATT | Died |
| Aliabadi et al 2009 | USA | 14/M  Immigrant | NA | NA | Headache  Altered consciousness | Multiple brain tuberculoma | NA | NA | Glucose = 70  mg/dl  Proteins= 307  mg/dL  Cells= 500  cells/ml | NA | NA | Brain biopsy= caseating granuloma demonstrating tuberculous granuloma | Tuberculous meningitis  brain tuberculoma | ATT and corticosteroids | Improved |
| Takahashi et al 2008 | Japan | 46/M | Negative | 2 weeks | Fever  Headache  Paradoxical paraparesis | Intradural extramedullary tuberculoma | Spine cord lesion paradoxically after start of ATT | NA | Glucose = 19  mg/dl  Proteins= 372  mg/dL  Cells= 434  cells/ml  ADA= elevated | Pus from spinal cord abscess = *Mycobacterium tuberculosis* | NA | NA | Tuberculous meningitis  Intradural extramedullary tuberculoma | ATT  Surgical removal of abscess | Improved |
| Park et al 2008 | Korea | 66/F | Negative | 2 weeks | Fever  Backpain  Paraparesis | Intradural extramedullary tuberculoma in cervical cord  Miliary tuberculoma brain | Paradoxically developed Vertebral tuberculosis | NA | NA | PCR biopsied brain tissue= *Mycobacterium tuberculosis* | NA | Spinal cord biopsy and PCR= tuberculous granuloma with *Mycobacterium tuberculosis* | Spinal tuberculosis  Miliary tuberculoma brain | ATT  Spinal cord surgery | Improved |
| Noh et al 2008 | Korea | 46/M | Negative | 2 months | Fever  Altered sensorium | Miliary tuberculoma brain | NA | Multiple choroid tubercles | NA | Urine and CSF smear= *Mycobacterium tuberculosis* | NA | NA | Tuberculous meningitis with Miliary tuberculoma brain | ATT and corticosteroids | Improved |
| Marschall et al 2008 | Switzerland | 48/M | HIV positive  CD4 count= > 100/mm3 | NA | Fever  Altered sensorium  Osteoarticular  Tuberculosis  total left knee arthroplasty | Multiple infarcts | NA | NA | NA | CSF= *Mycobacterium tuberculosis* | MDR strain | Sample from knee lesions smear= *Mycobacterium tuberculosis*  Autopsy= Tuberculous meningitis | Tuberculous meningitis | ATT and corticosteroids | Died |
| Lunn et al 2008 | UK | 35/F | HIV positive  CD4 count= > 6/mm3 | 3 months | Progressive flaccid paraparesis  with radicular sensory loss | Multiple tuberculoma brain  Multiple tuberculoma spinal cord | NA | NA | Increased protein  Rest parameters normal | Sputum smear= *Mycobacterium tuberculosis* | NA | NA | Multiple tuberculoma brain  Multiple tuberculoma spinal cord | ATT and corticosteroids  ART | Improved |
| Lee et al 2008 | Korea | 52/F | NA | 3 weeks | Headache vomiting  Mental  Deterioration  Two weeks later akinetic mutism | Multiple intracranial tuberculoma  Anterior cerebral artery infarction | Paradoxically increased size of infarction, increased number of  tuberculomas, and worsening of cerebral arteritis | NA | NA | NA | NA | NA | Multiple intracranial tuberculoma  Anterior cerebral artery infarction | NA | NA |
| Ho and Hsu 2008 | Taiwan | 70/F | NA | 1 month | Akinetic mutism | Multiple intracranial tuberculoma | NA | NA | Lymphocytic pleocytosis | NA | NA | NA | Tuberculous meningitis with Miliary tuberculoma brain | ATT and corticosteroids | Improved |
| Blackmore et al 2008 | New Zealand | 42/F  Immigrant | NA | NA | Fever  Altered sensorium  Focal neurological deficits  Clinical deterioration after start of ATT | A large lesion in left parietal region | Paradoxical multiple new lesion in whole brain | NA | Glucose = normal  Proteins= 193  mg/dL  Cells= 216  cells/ml | Sputum smear= *Mycobacterium tuberculosis* | NA | Brain biopsy= tuberculous granuloma with presence of AFB | Tuberculous meningitis with tuberculoma brain | ATT and corticosteroids  Infliximab | Improved but significant disability persisted |
| Surani et al 2007 | USA | NA | NA | NA | Altered sensorium | Left parietal tuberculoma  Meningeal enhancement | NA | NA | NA | CSF culture= *Mycobacterium tuberculosis* | NA | NA | Tuberculous meningitis with tuberculoma brain | ATT | NA |
| Nateghian et al 2007 | USA | 5/F | NA | 1 month | Headache  Seizures | Miliary tuberculoma brain  A large conglomerated left parasagittal tuberculoma | NA | NA | NA | Gastric aspirate= *Mycobacterium tuberculosis* | NA | NA | Miliary tuberculoma brain | ATT and corticosteroids | Improved |
| Kyoung et al 2007 | Korea | 18/F | NA | 2 weeks | Fever  Headache  Dysnoea | Miliary tuberculoma brain  Tuberculoma in liver, spleen and kidney | NA | NA | Glucose = 41mg/dl  Proteins= 57  mg/dL  Cells= 300 mm3 | Urine culture= *Mycobacterium tuberculosis* | NA | Spleen biopsy= tuberculous granuloma | Tuberculous meningitis with tuberculoma brain | ATT | Improved |
| Korri and Awada 2007 | France | 49/M | NA | 15 days | Paraparesis | Intramedullary spinal cord tuberculoma at T12 | NA | NA | NA | Sputum smear= *Mycobacterium tuberculosis* | NA | NA | Intramedullary spinal cord tuberculoma | ATT | Improved |
| Ahn et al 2007 | South Korea | 23/F | NA | 2 months | Fever | Multiple intracranial tuberculoma | NA | NA | NA | NA | NA | Bronchoscopy biopsy= chronic granulomatous  inflammation with caseous | Multiple intracranial tuberculoma | ATT and corticosteroids | Improved |
| Alkhani et al 2006 | Saudi Arabia | 25/F | NA | 1 month | Headache  Coma | Miliary tuberculoma brain | NA | NA | Increased cells | NA | NA | Brain biopsy= inflammatory caseating  Granulomas  *Mycobacterium tuberculosis was cultured* | Miliary tuberculoma brain | ATT and corticosteroids | Improved |
| Uysal et al 2005 | Turkey | 5 months/M | NA | NA | Respiratory complaints | Asymptomatic multiple intracranial tuberculoma, largest in the pons | NA | NA | Normal | Gastric aspirate= *Mycobacterium tuberculosis* | NA | NA | Asymptomatic multiple intracranial tuberculoma | ATT | Improved |
| Innocenti et al 2005 | USA | 38/M | NA | 2 months | Fever,  headache  and vomiting  Focal neurological deficit | Multiple intracranial tuberculoma  Miliary lesions in liver | NA | NA | Increased cells and protein  PCR and  Culture= Mycobacterium tuberculosis | NA | NA | NA | Disseminated tuberculosis | ATT | Improved |
| Ceylan and Gencer 2005 | Turkey | 12/F | NA | 1 month | Headache  Seizures | Multiple intracranial tuberculoma | NA | NA | NA | Gastric aspirate and sputum smear= *Mycobacterium tuberculosis* | NA | NA | Multiple intracranial tuberculoma | ATT | Improved |
| Bas et al 2005 | Turkey | 13/F | NA | NA | Fever  Headache  ATT was started for miliary tuberculosis | Multiple intracranial tuberculoma | Symptoms aggravated  Size of tuberculoma increasing in size | NA | NA | Sputum smear= *Mycobacterium tuberculosis* | NA | Brain biopsy= tuberculous granuloma | Multiple intracranial tuberculoma | ATT and corticosteroids | Improved |
| Akritidis et al 2005 | Greece | 68/M | NA | 2 months | Fever  Headache  Mental deterioration | Multiple intracranial tuberculoma | NA | NA | Glucose = 56 mg/dl  Proteins= 74  mg/dL  Cells= 32 mm3 | BAL= *Mycobacterium tuberculosis* | NA | NA | Multiple intracranial tuberculoma | ATT and corticosteroids | Improved |
| Yen et al 2003 | Taiwan. | 67/M | NA | 3 weeks | Fever  ATT was started for miliary tuberculosis  After two months he developed thoracic transverse myelopathy | NA | Paradoxical appearance of miliary intracranial tuberculoma  Multiple intramedullary tuberculoma spinal cord | NA | Glucose = 68 mg/dl  Proteins= 80  mg/dL  Cells= 05 mm3 | NA | NA | The pleural biopsy= tuberculous granuloma  Biopsy of excised spinal lesion= tuberculous granuloma. PCR detected *Mycobacterium tuberculosis* | Miliary intracranial tuberculoma  Multiple intramedullary tuberculoma spinal cord | ATT  Surgical excision of spinal cord lesion | Improved |
| Gasparetto et al 2003 | Brazil | 17/F  Pregnant | NA | 7 days | Fever  Headache  Sudden onset of paraparesis | Multiple intracranial tuberculoma | NA | NA | NA | NA | NA | Brain biopsy= tuberculous granuloma with presence of AFB | Multiple intracranial tuberculoma | ATT and corticosteroids | Improved |
| Liu et al 2002 | Taiwan | 26/F | Negative | 15 days | Fever  Headache  Paradoxical hemiparesis | Miliary tuberculoma brain | Paradoxical enlargement was noted | NA | Glucose = 38 mg/dl  Proteins= 591  mg/dL  Cells= 30 mm3 | Bone marrow biopsy= *Mycobacterium tuberculosis* | NA | NA | Tuberculous meningitis with tuberculoma brain | ATT and corticosteroids | Improved |
| Janner et al 2000 | USA | 8 months/M | NA | 3 weeks | Respiratory symptoms only | Multiple intracranial tuberculoma | NA | NA | Normal | Gastric aspirate and sputum smear= *Mycobacterium tuberculosis* | NA | NA | Multiple intracranial tuberculoma | ATT | Improved |
| Shinmura et al 1999  Autopsy based study | Japan | 65/F | End-stage renal disease | NA | Altered sensorium | Multiple intracranial tuberculoma | NA | NA | NA | Sputum smear= *Mycobacterium tuberculosis* | NA | Tuberculous lesions were seen in  liver, spleen,  heart, and lymph nodes  tuberculous granuloma with presence of AFB | Multiple intracranial tuberculoma | NA | NA |
| Nishikage et al 1999  Autopsy based study | Japan | 65/F | End-stage renal disease | NA | Fever  Altered sensorium | NA | NA | NA | NA | Sputum culture = *Mycobacterium tuberculosis* | NA | Autopsy= tuberculous granuloma in brain and lungs | Multiple intracranial tuberculoma | NA | NA |
| Jackman 1999 | USA | 9 months/F | NA | 1 month | Fever  Altered sensorium | NA | NA | NA | Glucose = 44 mg/dl  Proteins= 63  mg/dL  Cells= 122 mm3 | CSF  culture = *Mycobacterium tuberculosis* | NA | NA | Tuberculous meningitis | ATT and corticosteroids | Improved |
| Muin and Zurin 1998 | Malaysia | 19/F | NA | 2 months | Fever  Headache  Vision loss  Bilateral papilloedema | Multiple large intracranial tuberculoma | NA | NA | NA | NA | NA | Brain biopsy= tuberculous granuloma with presence of AFB | Multiple intracranial tuberculoma | ATT and corticosteroids | Improved |
|  |  | 37/M | NA | 5 months | Fever  Headache  Vision loss  Bilateral papilloedema | Multiple large intracranial tuberculoma in posterior fossa | NA | NA | NA | NA | NA | Brain biopsy= tuberculous granuloma with presence of AFB | Multiple intracranial tuberculoma | ATT and corticosteroids | Improved |
| Crump et al 1998 | UK | 35/M | HIV positive  (CD4 T cell count, 210/mm3)  On ART | 2 weeks | Lymphadenopathy  Patient received ATT  5 months after ATT, he had seizures | NA | Paradoxically developed multiple large intracranial tuberculoma | NA | NA | NA | NA | Lymph node biopsy= tuberculous granuloma with presence of AFB  biopsy= tuberculous granuloma with presence of AFB | Multiple intracranial tuberculoma | ATT and corticosteroids | Improved |
| Reiser et al 1997 | Germany | 33/F  Immigrant | NA | 2 months | Fever  Headache  Altered consciousness  Stared ATT and symptoms improved | NA | Despite improvement in symptoms, he developed multiple tuberculoma brain | NA | Glucose = 18 mg/dl  Proteins= 118  mg/dL  Cells= 257 mm3 | CSF  PCR = *Mycobacterium tuberculosis* | NA | NA | Tuberculous meningitis with tuberculoma brain | ATT and corticosteroids | Improved |
| Ajay et al 1996 | India | 8/F | NA | 2 weeks | Fever  Patient took ATT for 9 months for miliary tuberculosis  She had Headache  Ataxia  Focal neurological deficit | NA | Despite improvement in symptoms, he developed multiple tuberculoma brain and hydrocephalus | NA | NA | NA | NA | Brain biopsy= tuberculous granuloma | Multiple intracranial tuberculoma | VP shunt  Surgical excision of granuloma  ATT | Improved |
| Ghosh and Senapati 1994 | India | 4 months/M | NA | 3 weeks | Respiratory symptoms  Seizures | NA | NA | NA | Glucose = 25 mg/dl  Proteins= 120  mg/dL  Cells=60 mm3 | CSF  smear = *Mycobacterium tuberculosis* | NA | NA | Tuberculous meningitis | ATT and corticosteroids | Improved |
| Geissmann and Minamoto 1994 | USA | 31/F | HIV infection  CD4 T cell count= 6/mm3 | 2 weeks | Headache  Seizures  Altered consciousness  Cervical lymphadenopathy  Patient received ATT | NA | Paradoxically developed brain tuberculoma | NA | Glucose = 51 mg/dl  Proteins= 68  mg/dL  Cells=11 mm3 | Sputum smear = *Mycobacterium tuberculosis* | NA | Lymph node biopsy= *Mycobacterium tuberculosis*    Brain biopsy= tuberculous granuloma | Tuberculous meningitis  Large intracranial tuberculomas | ATT and corticosteroids | Died |
| Cilow et al 1994 | USA | 31/M  Immigrant | NA | 10 days | Paraparesis after start of ATT for miliary tuberculosis | NA | Paradoxically developed intramedullary spinal cord tuberculoma | NA | Glucose = 40 mg/dl  Proteins= 91  mg/dL  Cells=12 mm3 | CSF  smear = *Mycobacterium tuberculosis* | NA | Biopsy of resected lesion = tuberculous granuloma | Spinal cord tuberculoma | Because of worsening tuberculoma was surgically removed  ATT and corticosteroids | Improved |
| Afghani and Lieberman 1994 | USA | 2/F | NA | 3 days | Fever  Respiratory symptoms  Altered sensorium  ATT was given | NA | Paradoxically developed meningeal enhancement | NA | Glucose = 41 mg/dl  Proteins= 56  mg/dL  Cells=32 mm3 | BAL culture= *Mycobacterium tuberculosis* | NA | NA | Tuberculous meningitis | ATT and corticosteroids | Improved |
| Shen et al 1993 | China | 30/M | NA | 1 week | Fever  Paraparesis | Intramedullary spinal cord tuberculomas in cervical and thoracic regions  Miliary brain tuberculomas | NA | NA | NA | NA | NA | NA | Miliary tuberculoma brain and spinal cord | ATT and corticosteroids | Improved |
| Gee et al 1992 | USA | 1/F | NA | NA | Patient was on ATT for 3 months for treatment of tuberculous meningitis  Suddenly, she had fever, seizures and hemiparesis | NA | Paradoxically developed miliary brain lesions  Infarct in right parieto-occipital region | NA | Lymphocytic pleocytosis | NA | NA | NA | Miliary tuberculoma brain | ATT | NA |
| Shibolet et al 1979 | Israel | 21/F  Pregnant | NA | NA | Patient was on ATT for treatment of tuberculous meningitis  After 6 months, soon after delivery patient had fever and headache and seizures | Hydrocephalus, for that VP shunt was inserted | After few months, she had paradoxical miliary tuberculosis | NA | Lymphocytic pleocytosis | Culture from tip of shunt= *Mycobacterium tuberculosis* | NA | NA | Miliary tuberculosis | ATT and shunt surgery | Died |
| Lieberman et al 1970 | Japan | 28/M | NA | NA | Headache  Hemiparesis  Diplopia | NA | NA | NA | Protein was mildly raised | Sputum culture = *Mycobacterium* | NA | Brain biopsy= tuberculous granuloma with presence of AFB | A large tuberculoma brain | ATT and corticosteroids  Surgical removal of tuberculoma | Improved |
| Case records of the Massachusetts General Hospital 1953 | USA | 46/F | NA | NA | Fever  Headache  Altered consciousness  Flaccid Paraparesis | NA | NA | NA | Glucose = NA  Proteins= 112  mg/dL  Cells=115 mm3 | NA | NA | Uterine biopsy= tuberculoid granuloma | Tuberculous meningitis  Tuberculous myelitis | NA | NA |
| Tooke 1935 | Canada | 1/M | NA | NA | Fever  Seizures  Marked respiratory symptoms | NA | NA | Choroid tubercles present | Few cells only | Sputum smear = *Mycobacterium* | NA | Autopsy= tuberculous involvement of many body parts including brain | Tuberculous meningitis | NA | Died |
| Rodin and Dickey 1928 | USA | 5/F | NA | NA | Fever | NA | NA | Choroid tubercles present | Markedly increasing cells | NA | NA | A guinea-pig inoculation test revealed AFB | Tuberculous meningitis | ATT | Died |

Greenberg J, Huertas-Arias B, Beltramo F. Miliary tuberculosis presenting as altered mental status. Consultant.

Published online March 4, 2021. doi:10.25270/con.2021.03.00005.

Azeem A, Ahmad F, Velagapudi M. Infliximab for Immune Reconstitution Inflammatory Syndrome (IRIS) in Tuberculous Meningitis; A Treatment Paradox. Open Forum Infectious Diseases. 2021;8(SUPPL 1). DOI: 10.1093/ofid/ofab466.1593.

Velásquez-Rimachi V, Orellana-Tovar I, Rodriguez-López E, López-Saavedra A, Esteban-Arias D, Pacheco-Barrios K, et al. Multiple tuberculomas in an immunocompetent patient and their diagnostic challenge in a high prevalence country: Case report and literature review. Indian Journal of Tuberculosis. 2020;67(3):286-94. DOI: 10.1016/j.ijtb.2020.05.007.

Vasconcelos G, Santos L, Couto C, Cruz M, Castro A. Miliary Brain Tuberculomas and Meningitis: Tuberculosis Beyond the Lungs. Eur J Case Rep Intern Med. 2020;7(12):001931. DOI: 10.12890/2020_001931.

Tetsuka S, Suzuki T, Ogawa T, Hashimoto R, Kato H. Central nervous system tuberculoma with miliary tuberculosis in the elderly. IDCases. 2020;19. DOI: 10.1016/j.idcr.2020.e00710.

Sivakami A, Shah H, Revati N. Multiple tuberculoma and choroid tubercles in a child with miliary tuberculosis. Journal of the Pediatrics Association of India. 2020;9(1):38-40. DOI:

10.4103/jpai.jpai_17_20.

Rahman N, Kumar D, Hampannavar MS, Jain A, Pannu AK. Classic miliary TB. QJM: An International Journal of Medicine. 2020;113(7):504-5. DOI: 10.1093/qjmed/hcz270.

Meriem M, Najla M, Nidhal B, Mejda B, Nizar L, Imen Y. Disseminated tuberculosis in a pregnant immunocompetent healthcare worker. Egyptian Journal of Chest Diseases and Tuberculosis. 2020;69(4):753-6. DOI: 10.4103/ejcdt.ejcdt-123-19.

Ish S, Rathi V, Ish P, Garkoti H, Sharma D. A rare initial presentation of miliary tuberculosis. Advances in Respiratory Medicine. 2020;88(5):462-3. DOI: 10.5603/ARM.a2020.0127.

Esposito SB, Levi J, Matuzsan ZM, Amaducci AM, Richardson DM. A case report of widely disseminated tuberculosis in immunocompetent adult male. Clinical Practice and Cases in Emergency Medicine. 2020;4(3):375. DOI: 10.5811/cpcem.2020.3.46183.

St Cyr G, Starke JR. Multiple Cranial Tuberculomas Without Meningitis in Two Infants With Miliary Tuberculosis. Pediatr Infect Dis J. 2019;38(12). DOI: 10.1097/inf.0000000000002464.

Sánchez-Códez MI, Lubián-Gutiérrez M, Fernández-Bravo C, Ley-Martos M. Pediatric miliary tuberculosis presenting with stroke: contribution to the paper “Tuberculosis of the central nervous system in children”. Child's Nervous System. 2019;35:1273-5. https://doi.org/10.1007/s00381-019-04270-5.

Hansen KC, Jensen-Fangel S, Hønge BL. Contribution to differential diagnosis of sarcoidosis and disseminated tuberculosis. BMJ Case Reports. 2019;12(11). DOI: 10.1136/bcr-2019-230652.

Boubnan Y, Lezrek O, Laghmari M, Cherkaoui O. Choroidal granuloma secondary to tuberculosis in an immunocompromised patient. Journal Francais d'Ophtalmologie. 2019;42(6). DOI: 10.1016/j.jfo.2018.11.021.

Amin S, Stone D, Anderlind C. A 32-Year-Old Woman With Miscarriage, Headache, Hepatitis, and Pulmonary Disease. Chest. 2019;155(4). DOI: 10.1016/j.chest.2018.10.001.

Zhan Y, Li B, Huo Y, Lin A, Wu H. A case of multiple organ tuberculosis. Radiology of Infectious Diseases. 2018;5(1):50-4. https://doi.org/10.1016/j.jrid.2018.02.003.

Vélez-Tirado N, Cote-Orozco JE. An Infectious Masquerader. Pediatr Neurol. 2018;81:51. DOI: 10.1016/j.pediatrneurol.2017.12.004.

Sharma R, Saini AG, Katoch D, Bhatia V. Never Forget the Optic Fundi in Tuberculosis! J Pediatr. 2018;195:305. DOI: 10.1016/j.jpeds.2017.11.035.

Sarswat S, Sachan D. Intraventricular tuberculoma: An unusual presentation of brain tuberculosis. Indian Journal of Tuberculosis. 2018;65(2):180-1. DOI: 10.1016/j.ijtb.2017.08.003.

Parija S, Lalitha CS, Naik S. Weber syndrome secondary to brain stem tuberculoma. Indian J Ophthalmol. 2018;66(7):1036-9. DOI: 10.4103/ijo.IJO_1040_17.

Meregildo Rodriguez ED. Central nervous system tuberculosis following delayed and initially missed lung miliary tuberculosis: a case report. Infez Med. 2018;26(3):270-5. https://www.infezmed.it/media/journal/Vol_26_3_2018_13.pdf.

Macauley P, Rapp M, Park S, Lamikanra O, Sharma P, Marcelin M, Sharma K. Miliary Tuberculosis Presenting With Meningitis in a Patient Treated With Mycophenolate for Lupus Nephritis: Challenges in Diagnosis and Review of the Literature. Journal of Investigative Medicine High Impact Case Reports. 2018;6. DOI: 10.1177/2324709618770226.

Ko Y, Mo E-K, Park YB, Kang M-R, Bae JS, Kim Y. Miliary tuberculosis mimicking brain metastasis from renal cell carcinoma. Journal of Neurocritical Care. 2018;11(1):47-53. https://doi.org/10.18700/jnc.180045.

Kim Y, Kim SP, Han S. Multiple tuberculomas invading the central nervous system as a paradoxical reaction in a kidney transplantation recipient. Saudi J Kidney Dis Transpl. 2018;29(3):719-22. DOI: 10.4103/1319-2442.235190.

Balal M, Ulu AC, Demirkıran M. Miliary Tuberculosis with Postpartum Involvement of the Central Nervous System. Turk J Neurol. 2018;24:273-4. DOI:10.4274/tnd.17093.

Pal S, Gupta A, Dhasmana R, Saini M, Khanduri R, Bhat N. Miliary tuberculosis in an immunocompetent adolescent. Indian Journal of Child Health. 2017;4(3):459-61. Doi: 10.32677/IJCH.2017.v04.i03.047.

Paez Soria E, Magnano P, Schlaen A, Luvini P, Arevalo Calderon G, Martinez Cartier M, et al. Tuberculous subretinal abscess in a non-HIV patient with miliary tuberculosis. Case Reports in Ophthalmology. 2017;7(3):570-8. DOI: 10.1159/000453447.

Madhyastha SP, Gopalaswamy V, Acharya RV, Doddamani A. Disseminated tuberculosis in relatively asymptomatic young woman. BMJ Case Reports. 2017;2017. doi:10.1136/bcr-2017-219276.

Guerrero-Laleona C, Calzada-Hernández J, Bustillo-Alonso M, Gil-Albarova J, Medrano-San Ildefonso M, Iglesias-Jiménez E, Noguera-Julian A. Disseminated Tuberculosis Resulting from Reinfection in a Pediatric Patient Sequentially Treated with Etanercept and Adalimumab. Pediatric Infectious Disease Journal. 2017;36(1):109-10. DOI: 10.1097/INF.0000000000001360.

Garg RK, Malhotra H, Kumar N, Ingole R, Pandey S. Simultaneous miliary lesions of brain and lungs: A diagnostic challenge. Annals of Indian Academy of Neurology. 2017;20(4):431-2. DOI: 10.4103/aian.AIAN_261_17.

Emamzadehfard S, Paydary K, Nabavizadeh SA. Anterior cerebral artery vasculopathy secondary to miliary TB. Applied Radiology. 2017;46(12):16-7. https://www.proquest.com/openview/c886a51ac8cacf9a956bc5b91e88339f/1?pq-origsite=gscholar&cbl=32662.

Zhao Y, Bu H, He JY. Intracranial miliary tuberculomas. QJM. 2016;109(1):65-6. DOI: 10.1093/qjmed/hcv156.

Viel-Thériault I, Thibeault R, Boucher FD, Drolet JP. Thalidomide in Refractory Tuberculomas and Pseudoabscesses. Pediatric Infectious Disease Journal. 2016;35(11):1262-4. DOI: 10.1097/INF.0000000000001285.

Rali P, Arshad H, Bihler E. A case of tuberculous meningitis with tuberculoma in nonimmunocompromised immigrant. Case reports in pulmonology. 2016;2016. http://dx.doi.org/10.1155/2016/9016142

Parmaksiz ET, Caglayan B, Kiral N, Dogan C, Salepci B, Comert S. An unusual case of multidrug resistant miliary tuberculosis. Archives of Clinical Infectious Diseases. 2016;11(4). doi: 10.5812/archcid.37805.

Baudel JL, Dubée V, Palle J, Ait-Oufella H. Seizures, Paraplegia, and Cough Unveiling Disseminated Tuberculosis. Am J Med. 2016;129(1). DOI: 10.1016/j.amjmed.2015.06.048.

Wang M-H, Liu X, Shen B. Disseminated tuberculosis in a patient taking anti-TNF therapy for Crohn's disease. ACG Case Reports Journal. 2015;3(1):45-8. doi:10.14309/crj.2015.97.

Tanaka T, Sekine A, Tsunoda Y, Takoi H, Lin SY, Yatagai Y, et al. Central nervous system manifestations of tuberculosis-associated immune reconstitution inflammatory syndrome during adalimumab therapy: A case report and review of the literature. Internal Medicine. 2015;54(7):847-51. DOI: 10.2169/internalmedicine.54.2828.

Prasad S, Varma M, Vidyasagar S. Concurrent intracerebral and intramedullary spinal tuberculomas in an immunocompetent individual. BMJ Case Reports. 2015;2015. DOI: 10.1136/bcr-2014-205496.

Islam MF, Mukherjee D, Kundu R, Niyogi PC, Das J. A case of disseminated tuberculosis with ocular involvement. Journal of Health and Allied Sciences NU. 2015;5(01):094-6. DOI: 10.1055/s-0040-1703875.

Hsieh HY, Chen LA. Multiple Intracranial Tuberculomas in a Patient With Noonan Syndrome. Acta Neurol Taiwan. 2015;24(2):71-2. http://www.ant-tnsjournal.com/Mag_Files/24-2/008.pdf.

Gunbatar H, Bulut G, Soyoral Y, Sertogullarindan B, Ebinc S. Presented with central nervous system involvement, extra- pulmonary and intra-pulmonary tuberculosis combined infection in a dialysis patient. Eastern Journal of Medicine. 2015;20(3):171-4. https://jag.journalagent.com/ejm/pdfs/EJM_20_3_171_174.pdf.

Gamell A, Ntamatungiro AJ, Battegay M, Letang E. Case Report: Disseminated tuberculosis in an HIV-infected child: rifampicin resistance detected by GeneXpert in a lymph node aspirate but not in cerebrospinal fluid. BMJ Case Reports. 2015;2015. doi:10.1136/bcr-2014-207997.

Diguvinti S, Damam S, Ubara KK, Dara C. Concurrent occurrence of both intracranial and intramedullary tuberculomas. Neuroimmunology and Neuroinflammation. 2015;2:118-20. DOI:10.4103/2347-8659.153980.

Arora S, Narang GS, Dhillon PK. Neurological Worsening in a Child of Miliary Tuberculosis with Neuro-Tuberculosis on Anti Tubercular Treatment. Journal of Nepal Paediatric Society. 2015;35(2). doi: http://dx.doi.org/10.3126/jnps.v35i2.10406.

Yoo KM, Joo EJ, Yeom JS, Chae SW, Lee SY, Han KJ. Dissemination of multidrug-resistant tuberculosis in a patient with acute HIV infection. BMC Infect Dis. 2014;14:462. doi: http://dx.doi.org/10.3126/jnps.v35i2.10406.

Tsai F-F, Shiao C-C, Lin S-Y, Tai H-M. Intracranial tuberculoma. QJM: An International Journal of Medicine. 2014;107(5):391-2. doi:10.1093/qjmed/hct158.

Ranganath S, Wilson B, Narasimhan A, Midturi JK. Disseminated Tuberculosis in an immunocompetent patient. Central European Journal of Medicine. 2014;9(1):144-7. DOI: 10.2478/s11536-013-0261-7.

Kumar S, Kumar R, Radotra BD, Singh M. Tubercular ventriculitis: an uncommon entity. Indian J Pediatr. 2014;81(6):608-10. DOI: 10.2478/s11536-013-0261-7.

Kenzaka T, Noda A. Cerebral tuberculoma. QJM. 2014;107(1):81. DOI: 10.1093/qjmed/hct010.

Iqbal N, Natarajan N, Periyasamy S, George S, Basheer A, Mookkappan S. Miliary tuberculosis with left brachial monoplegia: A case report. The Australasian Medical Journal. 2014;7(10):400. http//dx.doi.org/10.4066/AMJ.2014.2169,

Hilal T, Hurley P, McCormick M. Disseminated tuberculosis with tuberculous meningitis in an immunocompetent host. Oxford Medical Case Reports. 2014;2014(7):125-8. doi:10.1093/omcr/omu049.

Sönmez A, Fatih Yetkin M, Köseoǧlu E, Mirza M. Multiple tuberculoma involving the brain and spinal cord in a patient with miliary pulmonary tuberculosis. Turk Noroloji Dergisi. 2013;19(4):151-2. DOI: 10.4274/Tnd.24445.

Smith BB, Hazelton BJ, Heywood AE, Snelling TL, Peacock KM, Macartney KK. Disseminated tuberculosis and tuberculous meningitis in Australian-born children; case reports and review of current epidemiology and management. J Paediatr Child Health. 2013;49(3). DOI: 10.1111/jpc.12035.

Patel NH, Sathvara P, Patel J, Vaghela D. Disseminated tuberculosis with paradoxical miliary tuberculomas of brain in a child with rickets. Journal of Pediatric Neurosciences. 2013;8(3):228-31. doi:10.4103/1817-1745.123687: 10.4103/1817-1745.123687.

Pasticci MB, Paciaroni M, Floridi P, Cecchini E, Baldelli F. Stroke in a patient with tuberculous meningitis and HIV infection. Mediterranean Journal of Hematology and Infectious Diseases. 2013;5(1). DOI: 10.4084/mjhid.2013.017.

Ishiwada N, Tokunaga O, Nagasawa K, Ichimoto K, Kinoshita K, Hishiki H, Kohno Y. Isoniazid-and streptomycin-resistant miliary tuberculosis complicated by intracranial tuberculoma in a Japanese infant. The Tohoku Journal of Experimental Medicine. 2013;229(3):221-5. https://doi.org/10.1620/tjem.229.221.

De La Riva P, Urtasun M, Castillo-Trivino T, Camino X, Arruti M, Mondragón E, De Munain AL. Clinical response to thalidomide in the treatment of intracranial tuberculomas: Case report. Clinical Neuropharmacology. 2013;36(2):70-2. DOI: 10.1097/WNF.0b013e318285caa1.

Winklhofer S, Kollias S. Incidental MRI finding of a pons tuberculoma in a patient with so-far-undiagnosed multisystemic tuberculosis infection. Clinical Imaging. 2012;36(5):623-5. DOI: 10.1016/j.clinimag.2011.11.013.

Verma R, Patil TB, Lalla R. Disseminated tuberculosis manifesting as pulmonary, meningeal and spinal tuberculosis in an immunocompetent patient. BMJ Case Reports. 2012. DOI: 10.1136/bcr-2012-007778.

Sundaram SS, Vijeratnam D, Mani R, Gibson D, Chauhan AJ. Tuberculous syringomyelia in an HIV-infected patient: A case report. International Journal of STD and AIDS. 2012;23(2):140-2. DOI: 10.1258/ijsa.2011.011104.

Şen A, Tuǧcu B, Soysal A, Yüksel B, Kocabiyik N, Arpaci B. Choroidal tuberculoma in two cases with multiple intracranial tuberculomas. Journal of Neurological Sciences. 2012;28(4):609-13. 28:(4)# 29;609-613, 2011. http://www.jns.dergisi.org/text.php3?id=474.

Morioka H, Matsumoto S, Kojima E, Takada K, Iwata S, Okachi S. Paradoxical infarct in tuberculous meningitis: a case report. Intern Med. 2012;51(8):949-51. DOI: 10.2169/internalmedicine.51.6830.

Jorge JH, Graciela C, Pablo AP, Luis SH. A life-threatening central nervous system-tuberculosis inflammatory reaction nonresponsive to corticosteroids and successfully controlled by infliximab in a young patient with a variant of juvenile idiopathic arthritis. J Clin Rheumatol. 2012;18(4):189-91. DOI: 10.2169/internalmedicine.51.6830.

Gönen I, ÖZşahin M, Yildirim M, Özdemir D, Yilmaz Aydin L, Kutlucan A, et al. Disseminated tuberculosis during the course of behçet's disease: A case report. Turkish Journal of Rheumatology. 2012;27(1):70-3. DOI: 10.5606/tjr.2012.010.

Das A, Das S, Mandal A, Halder A. Cerebral tuberculoma as a manifestation of paradoxical reaction in patients with pulmonary and extrapulmonary tuberculosis. Journal of Neurosciences in Rural Practice. 2012;3(3):350-4. DOI: 10.4103/0976-3147.102622.

Chaudhry LA, Ebtesam B-E, Al-Solaiman S. Milliary tuberculosis with unusual paradoxical response at 3 weeks of antituberculous treatment. J Coll Physicians Surg Pak. 2012;22(1):43-5. https://www.jcpsp.pk/archive/2012/Jan2012/12.pdf.

Yasar KK, Pehlivanoglu F, Sengoz G, Ince ER, Sandikci S. Tuberculous meningoencephalitis with severe neurological sequel in an immigrant child. Journal of Neurosciences in Rural Practice. 2011;2(1):77-9. DOI: 10.4103/0976-3147.80114.

Yasar KK, Pehlivanoglu F, Sengoz G, Ayrancioglu N. A case of tuberculous meningitis with multiple intracranial tuberculomas and miliary tuberculosis and choroid tubercles. Infection. 2011;39(4):395-6. DOI: 10.1007/s15010-011-0144-2.

Undrakonda V, Umakanth S. Short duration respiratory illness with abducens palsy in a young man. BMJ Case Rep. 2011;2011. DOI: 10.1136/bcr.07.2011.4470.

Papastefanou VP, Cohen VM. Tuberculoma of the choroid masquerading as a choroidal melanoma. Eye. 2011;25(11):1519-20. DOI: 10.1038/eye.2011.205.

Padhi TR, Basu S, Das T, Samal B. Optic disc tuberculoma in a patient with miliary tuberculosis. Ocular Immunology And Inflammation. 2011;19(1):67-8. DOI: 10.3109/09273948.2010.515374.

Ikegame S, Wakamatsu K, Fujita M, Nakanishi Y, Harada M, Kajiki A. A case of isoniazid-resistant miliary tuberculosis in which tuberculous meningitis paradoxically developed despite systemic improvement. J Infect Chemother. 2011;17(5):689-93. DOI: 10.1007/s10156-011-0218-1.

Hess S, Hospach T, Nossal R, Dannecker G, Magdorf K, Uhlemann F. Life-threatening disseminated tuberculosis as a complication of TNF-α blockade in an adolescent. European journal of pediatrics. 2011;170:1337-42. DOI 10.1007/s00431-011-1501-y.

Seif F, Armitage K, Petrozzi M. Unusual presentation of a common disease: disseminated tuberculosis in an immunocompetent patient. The American Journal of Medicine. 2010;123(9). doi:10.1016/j.amjmed.2010.01.021.

Rey LC, Sousa AQ. Severe disseminated tuberculosis in a 4-year-old girl. Braz J Infect Dis. 2010;14(6):639-40. DOI: 10.1590/s1413-86702010000600016.

Ottaviani S, Meyer O, Dieudé P. Intramedullary tuberculoma during infliximab therapy. Rheumatology. 2010;49(1):42. doi:10.1093/rheumatology/kep267.

Krygowski JD, Brennen DF, Counselman FL. Intracranial tuberculomas: an unusual cause of altered mental status in a pediatric patient. J Emerg Med. 2010;38(3):323-7. DOI: 10.1016/j.jemermed.2007.10.060.

Aslan S, Gulsun S, Atalay B. Disseminated tuberculosis complicated with tuberculous meningitis, miliary tuberculosis, and thoracal bone fracture while investigating a cervical lymphadenopathy. Tuberculosis: a hidden enemy? Neurosciences Journal. 2010;15(2):129-30. https://nsj.org.sa/content/nsj/15/2/129.full.pdf.

Vishnubhotla S, Siddhartha B, Sivramakrishna G, Lakshmi A, Kumaraswamy R. Proptosis in a post‐renal transplant patient with disseminated tuberculosis. Transplant Infectious Disease. 2009;11(3):241-2. DOI: 10.1111/j.1399-3062.2009.00394.x.

Aliabadi H, Pradhan A, Nakaji P, Grant G, Fuchs H. Miliary tuberculosis presenting with neurological symptoms. Barrow Quarterly. 2008; 24: No 1. https://www.barrowneuro.org/wp-content/uploads/Miliary-Tuberculosis-Presenting-with-Neurological-Symptoms-1.pdf.

Takahashi H, Ito S, Kojima S, Tanno T, Hattori T. Intradural extramedullary tuberculoma of the thoracic spine: Paradoxical response to antituberculous therapy. Internal Medicine. 2008;47(8):797-8. DOI: 10.2169/internalmedicine.47.0839.

Park HS, Song YJ, Song YJ. Multiple tuberculoma involving the brain and spinal cord in a patient with miliary pulmonary tuberculosis. Journal of Korean Neurosurgical Society. 2008;44(1):36-9. DOI: 10.3340/jkns.2008.44.1.36.

Noh JY, Heo JY, Lee KG, Yoon YK, Lee J, Song JY, et al. A case of disseminated tuberculosis with miliary central nervous system tuberculoma. Infection and Chemotherapy. 2008;40(6):323-6. DOI : 10.3947/ic.2008.40.6.323.

Marschall J, Evison JM, Droz S, Studer UC, Zimmerli S. Disseminated tuberculosis following total knee arthroplasty in an HIV patient. Infection. 2008;36(3):274-8. DOI: 10.1007/s15010-007-7011-1.

Lunn MPT, Clarke C, Aldeen T. Acute paraplegia in a patient with AIDS and a normal CSF examination. Journal of Hospital Medicine. 2008;3(3):279-80. DOI: 10.1002/jhm.303.

Lee SI, Park JH, Kim JH. Paradoxical progression of intracranial tuberculomas and anterior cerebral artery infarction. Neurology. 2008;71(1):68. DOI: 10.1212/01.wnl.0000316309.86367.3e.

Ho BL, Hsu CY. Miliary intracranial tuberculomas presenting as rapidly reversible encephalopathy. Acta Neurol Taiwan. 2008;17(2):149-50. http://www.ant-tnsjournal.com/Mag_Files/17-2/dw2008626151338_17-2%20p149.pdf.

Blackmore TK, Manning L, Taylor WJ, Wallis RS. Therapeutic use of infliximab in tuberculosis to control severe paradoxical reaction of the brain and lymph nodes. Clin Infect Dis. 2008;47(10). DOI: 10.1086/592695.

Nateghian A, Robinson JL, Fanning A. A child with headache, cough, weight loss, and seizure. Clin Infect Dis. 2007;44(10):1341-2, 84-6. DOI: 10.1086/515400.

Kyoung TK, Dong JN, Sang HH, Sung SJ, Kyoung MM, Dong JK, et al. Unusual presentation of miliary tuberculosis. Tuberculosis and Respiratory Diseases. 2007;63(1):67-70. DOI: 10.4046/trd.2007.63.1.67.

Korri H, Awada A. Miliary tuberculosis and intramedullary tuberculoma of the conus medullaris. Revue Neurologique. 2007;163(11):1106-8. DOI: 10.1016/S0035-3787(07)74186-3.

Ahn JY, Chang JH, Kim KS, Kim WJ. Disseminated tuberculosis with multiple intracerebral tuberculomas in a patient with anorexia nervosa. Int J Eat Disord. 2007;40(3):288-91. DOI: 10.1002/eat.20368.

Alkhani A, Al-Otaibi F, Cupler EJ, Lach B. Miliary tuberculomas of the brain: case report. Clin Neurol Neurosurg. 2006;108(4):411-4. DOI: 10.1016/j.clineuro.2005.01.003.

Uysal G, Gursoy T, Altunc U, Guven A. Asymptomatic pons tuberculoma in an infant with military tuberculosis. Saudi Medical Journal. 2005;26(8):1277-9. https://nsj.org.sa/content/10/4/309.short.

Innocenti R, Degl'Innocenti L, Fronzaroli C, Ferrante F, Corradi F. Radiologic miliary patterns of cerebral tuberculosis. Archives of Neurology. 2005;62(1):153-4. DOI: 10.1001/archneur.62.1.153.

Ceylan E, Gencer M. Miliary tuberculosis associated with multiple intracranial tuberculomas. The Tohoku Journal of Experimental Medicine. 2005;205(4):367-70. https://doi.org/10.1620/tjem.205.367.

Bas NS, Güzey FK, Emel E, Alatas I, Sel B. Paradoxical intracranial tuberculoma requiring surgical treatment. Pediatr Neurosurg. 2005;41(4):201-5. DOI: 10.1159/000086562.

Akritidis N, Galiatsou E, Kakadellis J, Dimas K, Paparounas K. Brain tuberculomas due to miliary tuberculosis. Southern medical journal. 2005;98(1):111-4.

Yen HL, Lee RJ, Lin JW, Chen HJ. Multiple tuberculomas in the brain and spinal cord: a case report. Spine (Phila Pa 1976). 2003;28(23). DOI: 10.1097/01.Brs.0000099114.40764.46.

Gasparetto EL, Tazoniero P, de Carvalho Neto A. Disseminated tuberculosis in a pregnant woman presenting with numerous brain tuberculomas: case report. Arq Neuropsiquiatr. 2003;61(3b):855-8. DOI: 10.1590/s0004-282x2003000500028.

Liu SF, Wu HS, Lai YF. Miliary lung lesions and multiple intracranial tumors in a 26-year-old woman. Respiration. 2002;69(5):471-2. DOI: 10.1159/000064012.

Janner D, Kirk S, McLeary M. Cerebral tuberculosis without neurologic signs and with normal cerebrospinal fluid. The Pediatric infectious disease journal. 2000;19(8):763. DOI: 10.1097/00006454-200008000-00021.

Shinmura Y, Tsutsui Y, Nishikage H, Maeda M, Mokuno K, Hirose Y. Two autopsy cases of multiple cerebral tuberculomas. Neuropathology. 1999;19(4):404-9. DOI: 10.1046/j.1440-1789.1999.00260.x

Nishikage H, Kosugi T, Danbara A, Yamamoto J, Shinmura Y, Tsutsui Y. Autopsy case of miliary tuberculosis with cerebral involvement in renal failure. Clinical and experimental nephrology. 1999;3:212-7. DOI https://doi.org/10.1007/s101570050037.

Jackman GA. Radiological case of the month. Archives of Pediatrics and Adolescent Medicine. 1999;153(8):887-8. DOI: 10.1001/archpedi.153.8.887.

Muin IA, Zurin AR. Pulmonary miliary tuberculosis with multiple intracerebral tuberculous granulomas--report of two cases. Br J Neurosurg. 1998;12(6):585-7. DOI: 10.1080/02688699844501.

Crump JA, Tyrer MJ, Lloyd-Owen SJ, Han LY, Lipman MC, Johnson MA. Miliary tuberculosis with paradoxical expansion of intracranial tuberculomas complicating human immunodeficiency virus infection in a patient receiving highly active antiretroviral therapy. Clinical Infectious Diseases. 1998;26(4):1008-9. DOI: 10.1086/517636.

Reiser M, Fätkenheuer G, Diehl V. Paradoxical expansion of intracranial tuberculomas during chemotherapy. J Infect. 1997;35(1):88-90. DOI: 10.1016/s0163-4453(97)91241-x.

Ajay S, Lakhkar B, Bhaskaranand N. Intracranial tuberculoma manifesting during treatment. Indian pediatrics. 1996;33:231-2.

Ghosh JB, Senapati S. Tuberculous meningitis in early infancy. Indian Pediatr. 1994;31(12):1568-9.

Geissmann K, Minamoto GY. Paradoxical presentation of intracranial tuberculomas after chemotherapy in a patient with aids. Clin Infect Dis. 1994;19(4):793-4. DOI: 10.1093/clinids/19.4.793.

Cilow JS, Ammirati M, Horwitz NH, Benzel EC. Intramedullary tuberculoma of the spinal cord: Case report. Neurosurgery. 1994;35(2):327-30.

Afghani B, Lieberman JM. Paradoxical enlargement or development of intracranial tuberculomas during therapy: case report and review. Clin Infect Dis. 1994;19(6):1092-9. DOI: 10.1093/clinids/19.6.1092.

Shen WC, Cheng TY, Lee SK, Ho YJ, Lee KR. Disseminated tuberculomas in spinal cord and brain demonstrated by MRI with gadolinium-DTPA. Neuroradiology. 1993;35(3):213-5. DOI: 10.1007/bf00588498.

Gee GT, Bazan Iii C, Jinkins JR. Miliary tuberculosis involving the brain: MR findings. American Journal of Roentgenology. 1992;159(5):1075-6. DOI: 10.2214/ajr.159.5.1414778.

Shibolet S, Dan M, Jedwab M. Recurrent miliary tuberculosis secondary to infected ventriculoatrial shunt. Chest. 1979;76(3):328-30. DOI: 10.1378/chest.76.3.328.

Lieberman A, Dart L, Bennett R. Intracerebral tuberculoma. Case report. J Neurosurg. 1970;33(3):331-3. DOI: 10.3171/jns.1970.33.3.0331.

WEEKLY clinicopathological exercises; tuberculous meningitis, tuberculous endometritis, miliary tuberculosis. N Engl J Med. 1953;248(10):432-5. DOI: 10.1056/nejm197308162890711.

Tooke FT. Tuberculosis of the Choroid Associated with Generalized Miliary Tuberculosis. Transactions of the American Ophthalmological Society. 1935;33:201. DOI: 10.1136/bjo.20.1.23.

Rodin FH, Dickey LB. TUBERCLE OF THE CHOROID IN MILIARY TUBERCULOSIS: CASE REPORT. Cal West Med. 1928;28(6):807-9. PMID: 18740728.
