## Supplementary item-2 for "Central Nervous System Complications in Miliary Pulmonary Tuberculosis: A Systematic Review Of Published Case Reports And Case Series"

| **Reference** | **Does the patient represent the whole experience of the investigator** | **Was the exposure adequately ascertained?** | **Was the outcome adequately ascertained?** | **Were other alternative causes that may explain the observation ruled out?** | **Was there a challenge and/or re-challenge phenomenon?** | **Was there a dose-response effect?** | **Was follow-up long enough for outcomes to occur?** | **Is the case(s) described with sufficient details to allow to allow practitioners make inferences related to their own practice?** | **Score** |
| --- | --- | --- | --- | --- | --- | --- | --- | --- | --- |
| Mnaili and Bourazza 2024 | **Yes** | **Yes** | **Yes** | **Yes** | **NA** | **Yes** | **Yes** | **Yes** | **7** |
| Jayadhi Widyakusum et al 2024 | **Yes** | **No** | **Yes** | **Yes** | **NA** | **Yes** | **Yes** | **Yes** | **6** |
| Kamali et al 2024 | **Yes** | **Yes** | **Yes** | **Yes** | **NA** | **Yes** | **Yes** | **Yes** | **7** |
| Gama e al 2024 | **Yes** | **Yes** | **Yes** | **Yes** | **NA** | **Yes** | **Yes** | **Yes** | **7** |
| El Aggari *et al 2024* | **Yes** | **Yes** | **Yes** | **Yes** | **NA** | **Yes** | **Yes** | **Yes** | **7** |
| Ashizawa et al 2024 | **Yes** | **Yes** | **Yes** | **Yes** | **NA** | **Yes** | **Yes** | **Yes** | **7** |
| Tong et al 2023 | **Yes** | **Yes** | **Yes** | **Yes** | **NA** | **Yes** | **Yes** | **Yes** | **7** |
| Tavares-Júnior et al 2023 | **Yes** | **Yes** | **Yes** | **Yes** | **NA** | **Yes** | **Yes** | **Yes** | **7** |
| Subedi et al 2023 | **Yes** | **Yes** | **Yes** | **Yes** | **NA** | **Yes** | **Yes** | **Yes** | **7** |
| Samad et al 2023 | **Yes** | **Yes** | **Yes** | **Yes** | **NA** | **Yes** | **Yes** | **Yes** | **7** |
| Muraoka et al 2023 | **Yes** | **Yes** | **Yes** | **Yes** | **NA** | **Yes** | **Yes** | **Yes** | **7** |
| Mukherjee et al 2023 | **Yes** | **Yes** | **Yes** | **Yes** | **NA** | **Yes** | **Yes** | **Yes** | **7** |
| Labbad and Hamzeh 2023 | **Yes** | **Yes** | **Yes** | **Yes** | **NA** | **Yes** | **Yes** | **Yes** | **7** |
| Idrissa et al 2023 | **Yes** | **Yes** | **Yes** | **Yes** | **NA** | **Yes** | **Yes** | **Yes** | **7** |
| Ahmed et al 2023 | **Yes** | **Yes** | **Yes** | **Yes** | **NA** | **Yes** | **Yes** | **Yes** | **7** |
| Wautlet et al 2022 | **Yes** | **Yes** | **Yes** | **Yes** | **NA** | **Yes** | **Yes** | **Yes** | **7** |
| Ohgiya et al 2022 | **Yes** | **No** | **Yes** | **Yes** | **NA** | **NA** | **NA** | **Yes** | **4** |
| Letchuman et al 2022 | **Yes** | **Yes** | **Yes** | **Yes** | **NA** | **Yes** | **Yes** | **Yes** | **7** |
| Khan et al 2022 | **Yes** | **No** | **Yes** | **Yes** | **NA** | **NA** | **NA** | **Yes** | **4** |
| Hirayama et al 2022 | **Yes** | **Yes** | **Yes** | **Yes** | **NA** | **Yes** | **Yes** | **Yes** | **7** |
| Christian and Johnston 2022 | **Yes** | **Yes** | **Yes** | **Yes** | **NA** | **Yes** | **Yes** | **Yes** | **7** |
| Ashraf Talesh et al 2022 | **Yes** | **Yes** | **Yes** | **Yes** | **NA** | **Yes** | **Yes** | **Yes** | **7** |
| de Andrade et al 2022 | **Yes** | **Yes** | **Yes** | **Yes** | **NA** | **Yes** | **Yes** | **Yes** | **7** |
| Wong et al 2021 | **Yes** | **Yes** | **Yes** | **Yes** | **NA** | **Yes** | **Yes** | **Yes** | **7** |
| Soni et al 2021  Preprint | **Yes** | **Yes** | **Yes** | **Yes** | **NA** | **Yes** | **Yes** | **Yes** | **7** |
| Sekkat et al 2021 | **Yes** | **Yes** | **Yes** | **Yes** | **NA** | **Yes** | **Yes** | **Yes** | **7** |
| Sachan et al 2021 | **Yes** | **Yes** | **Yes** | **Yes** | **NA** | **Yes** | **Yes** | **Yes** | **7** |
| Reddy et al 2021 | **Yes** | **Yes** | **Yes** | **Yes** | **NA** | **Yes** | **Yes** | **Yes** | **7** |
| Pinzon et al 2021 | **Yes** | **Yes** | **Yes** | **Yes** | **NA** | **Yes** | **Yes** | **Yes** | **7** |
| Musa et al 2021 | **Yes** | **Yes** | **Yes** | **Yes** | **NA** | **Yes** | **Yes** | **Yes** | **7** |
| Kalita et al 2021 | **Yes** | **Yes** | **Yes** | **Yes** | **NA** | **Yes** | **Yes** | **Yes** | **7** |
| Duc et al 2021 | **Yes** | **Yes** | **Yes** | **Yes** | **NA** | **Yes** | **Yes** | **Yes** | **7** |
| Dormegnie et al 2021 | **Yes** | **Yes** | **Yes** | **Yes** | **NA** | **Yes** | **Yes** | **Yes** | **7** |
| Cheng et al 2021 | **Yes** | **Yes** | **Yes** | **Yes** | **NA** | **Yes** | **Yes** | **Yes** | **7** |
| Bouchentouf  2021 | **Yes** | **Yes** | **Yes** | **Yes** | **NA** | **Yes** | **Yes** | **Yes** | **7** |
| Benchekroun et al 2021 | **Yes** | **Yes** | **Yes** | **Yes** | **NA** | **Yes** | **Yes** | **Yes** | **7** |
| Bǎiceanu et al 2021 | **Yes** | **Yes** | **Yes** | **Yes** | **NA** | **Yes** | **Yes** | **Yes** | **7** |
| Greenberg e et al 2021 | **Yes** | **Yes** | **Yes** | **Yes** | **NA** | **Yes** | **Yes** | **Yes** | **7** |
| Velásquez-Rimachi et al 2020 | **Yes** | **No** | **Yes** | **Yes** | **NA** | **Yes** | **Yes** | **Yes** | **6** |
| Vasconcelos et al 2020 | **Yes** | **No** | **Yes** | **Yes** | **NA** | **NA** | **NA** | **Yes** | **4** |
| Tetsuka et al 2020 | **Yes** | **Yes** | **Yes** | **Yes** | **NA** | **Yes** | **Yes** | **Yes** | **7** |
| Sivakami et al 2020 | **Yes** | **Yes** | **Yes** | **Yes** | **NA** | **Yes** | **Yes** | **Yes** | **7** |
| Rahman et al 2020 | **Yes** | **Yes** | **Yes** | **Yes** | **NA** | **Yes** | **Yes** | **Yes** | **7** |
| Meriem et al 2020 | **Yes** | **Yes** | **Yes** | **Yes** | **NA** | **Yes** | **Yes** | **Yes** | **7** |
| Ish et al 2020 | **Yes** | **No** | **Yes** | **Yes** | **NA** | **NA** | **Yes** | **NA** | **5** |
| Esposito et al 2020 | **Yes** | **Yes** | **Yes** | **Yes** | **NA** | **Yes** | **Yes** | **Yes** | **7** |
| St Cyr and Starke 2020 | **Yes** | **Yes** | **Yes** | **Yes** | **NA** | **Yes** | **Yes** | **Yes** | **7** |
|  | **Yes** | **Yes** | **Yes** | **Yes** | **NA** | **Yes** | **Yes** | **Yes** | **7** |
| Sánchez-Códez et al 2019 | **Yes** | **No** | **Yes** | **Yes** | **NA** | **NA** | **NA** | **Yes** | **4** |
| Hansen et al 2019 | **Yes** | **Yes** | **Yes** | **Yes** | **NA** | **Yes** | **Yes** | **Yes** | **7** |
| Boubnan et al 2019 | **Yes** | **Yes** | **Yes** | **Yes** | **NA** | **Yes** | **Yes** | **Yes** | **7** |
| Amin et al 2019 | **Yes** | **Yes** | **Yes** | **Yes** | **NA** | **Yes** | **Yes** | **Yes** | **7** |
| Zhan et al 2018 | **Yes** | **Yes** | **Yes** | **Yes** | **NA** | **Yes** | **Yes** | **Yes** | **7** |
| Vélez-Tirado and Cote-Orozco 2018 | **Yes** | **Yes** | **Yes** | **Yes** | **NA** | **Yes** | **Yes** | **Yes** | **7** |
| Sharma et al 2018 | **Yes** | **Yes** | **Yes** | **Yes** | **NA** | **Yes** | **Yes** | **Yes** | **7** |
| Saraswat et al, 2018 | **Yes** | **No** | **Yes** | **Yes** | **NA** | **NA** | **Yes** | **NA** | **5** |
| Parija et al, 2018 | **Yes** | **Yes** | **Yes** | **Yes** | **NA** | **Yes** | **Yes** | **Yes** | **7** |
| Rodriguez et al, 2018 | **Yes** | **Yes** | **Yes** | **Yes** | **NA** | **Yes** | **Yes** | **Yes** | **7** |
| Macauley et al, 2018 | **Yes** | **Yes** | **Yes** | **Yes** | **NA** | **Yes** | **Yes** | **Yes** | **7** |
| Ko et al, 2018 | **Yes** | **Yes** | **Yes** | **Yes** | **NA** | **Yes** | **Yes** | **Yes** | **7** |
| Kim et al, 2018 | **Yes** | **Yes** | **Yes** | **Yes** | **NA** | **Yes** | **Yes** | **Yes** | **7** |
| Balal et al, 2018 | **Yes** | **No** | **Yes** | **Yes** | **NA** | **NA** | **Yes** | **NA** | **5** |
| Pal et al, 2017 | **Yes** | **Yes** | **Yes** | **Yes** | **NA** | **Yes** | **Yes** | **Yes** | **7** |
| Soria et al, 2017 | **Yes** | **No** | **Yes** | **Yes** | **NA** | **Yes** | **Yes** | **Yes** | **6** |
| Madhyastha et al, 2017 | **Yes** | **Yes** | **Yes** | **Yes** | **NA** | **Yes** | **Yes** | **Yes** | **7** |
| Laleona et al, 2017 | **Yes** | **Yes** | **Yes** | **Yes** | **NA** | **Yes** | **Yes** | **Yes** | **7** |
| Garg et al, 2017 | **Yes** | **Yes** | **Yes** | **Yes** | **NA** | **Yes** | **Yes** | **Yes** | **7** |
| Emamzadehfard et al, 2017 | **Yes** | **No** | **Yes** | **Yes** | **NA** | **NA** | **Yes** | **NA** | **5** |
| Zhao et al, 2016 | **Yes** | **Yes** | **Yes** | **Yes** | **NA** | **Yes** | **Yes** | **Yes** | **7** |
| Viel-Thériault et al, 2016 | **Yes** | **Yes** | **Yes** | **Yes** | **NA** | **Yes** | **Yes** | **Yes** | **7** |
| Rali et al, 2016 | **Yes** | **Yes** | **Yes** | **Yes** | **NA** | **Yes** | **Yes** | **Yes** | **7** |
| Parmaksiz et al, 2016 | **Yes** | **Yes** | **Yes** | **Yes** | **NA** | **Yes** | **Yes** | **Yes** | **7** |
| Baudel et al, 2015 | **Yes** | **Yes** | **Yes** | **Yes** | **NA** | **Yes** | **Yes** | **Yes** | **7** |
| Wang et al, 2015 | **Yes** | **Yes** | **Yes** | **Yes** | **NA** | **Yes** | **Yes** | **Yes** | **7** |
| Tanaka et al, 2015 | **Yes** | **Yes** | **Yes** | **Yes** | **NA** | **Yes** | **Yes** | **Yes** | **7** |
| Prasad et al, 2015 | **Yes** | **Yes** | **Yes** | **Yes** | **NA** | **Yes** | **Yes** | **Yes** | **7** |
| Islam et al, 2015 | **Yes** | **Yes** | **Yes** | **Yes** | **NA** | **Yes** | **Yes** | **Yes** | **7** |
| Hsieh et al, 2015 | **Yes** | **Yes** | **Yes** | **Yes** | **NA** | **Yes** | **Yes** | **Yes** | **7** |
| Gunbatar et al, 2015 | **Yes** | **No** | **Yes** | **Yes** | **NA** | **NA** | **Yes** | **NA** | **5** |
| Gamell et al, 2015 | **Yes** | **Yes** | **Yes** | **Yes** | **NA** | **Yes** | **Yes** | **Yes** | **7** |
| Diguvinti et al, 2015 | **Yes** | **Yes** | **Yes** | **Yes** | **NA** | **Yes** | **Yes** | **Yes** | **7** |
| Arora et al, 2015 | **Yes** | **Yes** | **Yes** | **Yes** | **NA** | **Yes** | **Yes** | **Yes** | **7** |
| Yoo et al 2014 | **Yes** | **Yes** | **Yes** | **Yes** | **NA** | **Yes** | **Yes** | **Yes** | **7** |
| Tsai et al 2014 | **Yes** | **Yes** | **Yes** | **Yes** | **NA** | **Yes** | **Yes** | **Yes** | **7** |
| Ranganath et al 2014 | **Yes** | **Yes** | **Yes** | **Yes** | **NA** | **Yes** | **Yes** | **Yes** | **7** |
| Kumar et al 2014 | **Yes** | **Yes** | **Yes** | **Yes** | **NA** | **Yes** | **Yes** | **Yes** | **7** |
| Kenzaka and Noda 2014 | **Yes** | **Yes** | **Yes** | **Yes** | **NA** | **Yes** | **Yes** | **Yes** | **7** |
| Iqbal et al 2014 | **Yes** | **Yes** | **Yes** | **Yes** | **NA** | **Yes** | **Yes** | **Yes** | **7** |
| Hilal et al 2014 | **Yes** | **Yes** | **Yes** | **Yes** | **NA** | **Yes** | **Yes** | **Yes** | **7** |
| Sönmez et al 2013 | **Yes** | **Yes** | **Yes** | **Yes** | **NA** | **Yes** | **Yes** | **Yes** | **7** |
| Smith et al 2013 | **Yes** | **Yes** | **Yes** | **Yes** | **NA** | **Yes** | **Yes** | **Yes** | **7** |
| Patel et al 2013 | **Yes** | **Yes** | **Yes** | **Yes** | **NA** | **Yes** | **Yes** | **Yes** | **7** |
| Pasticci et al 2013 | **Yes** | **Yes** | **Yes** | **Yes** | **NA** | **Yes** | **Yes** | **Yes** | **7** |
| Ishiwada et al 2013 | **Yes** | **Yes** | **Yes** | **Yes** | **NA** | **Yes** | **Yes** | **Yes** | **7** |
| De La Riva et al 2013 | **Yes** | **Yes** | **Yes** | **Yes** | **NA** | **Yes** | **Yes** | **Yes** | **7** |
| Winklhofer and Kollias  2012 | **Yes** | **Yes** | **Yes** | **Yes** | **NA** | **Yes** | **Yes** | **Yes** | **7** |
| Verma et al 2012 | **Yes** | **Yes** | **Yes** | **Yes** | **NA** | **Yes** | **Yes** | **Yes** | **7** |
| Sundaram et al 2012 | **Yes** | **Yes** | **Yes** | **Yes** | **NA** | **Yes** | **Yes** | **Yes** | **7** |
| Şen et al 2012 | **Yes** | **No** | **Yes** | **Yes** | **NA** | **NA** | **Yes** | **NA** | **5** |
| Morioka et al 2012 | **Yes** | **Yes** | **Yes** | **Yes** | **NA** | **Yes** | **Yes** | **Yes** | **7** |
| Jorge et al 2012 | **Yes** | **Yes** | **Yes** | **Yes** | **NA** | **Yes** | **Yes** | **Yes** | **7** |
| Gönen et al 2012 | **Yes** | **Yes** | **Yes** | **Yes** | **NA** | **Yes** | **Yes** | **Yes** | **7** |
| Das et al 2012 | **Yes** | **Yes** | **Yes** | **Yes** | **NA** | **Yes** | **Yes** | **Yes** | **7** |
|  | **Yes** | **Yes** | **Yes** | **Yes** | **NA** | **Yes** | **Yes** | **Yes** | **7** |
| Chaudhry et al 2012 | **Yes** | **Yes** | **Yes** | **Yes** | **NA** | **Yes** | **Yes** | **Yes** | **7** |
| Yasar et al 2011 | **Yes** | **Yes** | **Yes** | **Yes** | **NA** | **Yes** | **Yes** | **Yes** | **7** |
| Yasar et al 2011 | **Yes** | **Yes** | **Yes** | **Yes** | **NA** | **Yes** | **Yes** | **Yes** | **7** |
| Undrakonda and Umakanth 2011 | **Yes** | **Yes** | **Yes** | **Yes** | **NA** | **Yes** | **Yes** | **Yes** | **7** |
| Papastefanou and Cohen 2011 | **Yes** | **Yes** | **Yes** | **Yes** | **NA** | **Yes** | **Yes** | **Yes** | **7** |
| Padhi et al 2011 | **Yes** | **Yes** | **Yes** | **Yes** | **NA** | **Yes** | **Yes** | **Yes** | **7** |
| Ikegame et al 2011 | **Yes** | **Yes** | **Yes** | **Yes** | **NA** | **Yes** | **Yes** | **Yes** | **7** |
| Hess et al 2011 | **Yes** | **Yes** | **Yes** | **Yes** | **NA** | **Yes** | **Yes** | **Yes** | **7** |
| Seif et al 2010 | **Yes** | **Yes** | **Yes** | **Yes** | **NA** | **Yes** | **Yes** | **Yes** | **7** |
| Rey and Sousa 2010 | **Yes** | **Yes** | **Yes** | **Yes** | **NA** | **Yes** | **Yes** | **Yes** | **7** |
| Ottaviani et al 2010 | **Yes** | **Yes** | **Yes** | **Yes** | **NA** | **Yes** | **Yes** | **Yes** | **7** |
| Krygowski et al 2010 | **Yes** | **Yes** | **Yes** | **Yes** | **NA** | **Yes** | **Yes** | **Yes** | **7** |
| Aslan et al 2010 | **Yes** | **Yes** | **Yes** | **Yes** | **NA** | **Yes** | **Yes** | **Yes** | **7** |
| Vishnubhotla et al 2009 | **Yes** | **Yes** | **Yes** | **Yes** | **NA** | **Yes** | **Yes** | **Yes** | **7** |
| Aliabadi et al 2009 | **Yes** | **Yes** | **Yes** | **Yes** | **NA** | **Yes** | **Yes** | **Yes** | **7** |
| Takahashi et al 2008 | **Yes** | **Yes** | **Yes** | **Yes** | **NA** | **Yes** | **Yes** | **Yes** | **7** |
| Park et al 2008 | **Yes** | **Yes** | **Yes** | **Yes** | **NA** | **Yes** | **Yes** | **Yes** | **7** |
| Noh et al 2008 | **Yes** | **Yes** | **Yes** | **Yes** | **NA** | **Yes** | **Yes** | **Yes** | **7** |
| Marschall et al 2008 | **Yes** | **Yes** | **Yes** | **Yes** | **NA** | **Yes** | **Yes** | **Yes** | **7** |
| Lunn et al 2008 | **Yes** | **Yes** | **Yes** | **Yes** | **NA** | **Yes** | **Yes** | **Yes** | **7** |
| Lee et al 2008 | **Yes** | **No** | **Yes** | **Yes** | **NA** | **NA** | **Yes** | **NA** | **5** |
| Ho and Hsu 2008 | **Yes** | **Yes** | **Yes** | **Yes** | **NA** | **Yes** | **Yes** | **Yes** | **7** |
| Blackmore et al 2008 | **Yes** | **Yes** | **Yes** | **Yes** | **NA** | **Yes** | **Yes** | **Yes** | **7** |
| Surani et al 2007 | **Yes** | **No** | **Yes** | **Yes** | **NA** | **NA** | **Yes** | **NA** | **5** |
| Nateghian et al 2007 | **Yes** | **Yes** | **Yes** | **Yes** | **NA** | **Yes** | **Yes** | **Yes** | **7** |
| Kyoung et al 2007 | **Yes** | **Yes** | **Yes** | **Yes** | **NA** | **Yes** | **Yes** | **Yes** | **7** |
| Korri and Awada 2007 | **Yes** | **Yes** | **Yes** | **Yes** | **NA** | **Yes** | **Yes** | **Yes** | **7** |
| Ahn et al 2007 | **Yes** | **Yes** | **Yes** | **Yes** | **NA** | **Yes** | **Yes** | **Yes** | **7** |
| Alkhani et al 2006 | **Yes** | **Yes** | **Yes** | **Yes** | **NA** | **Yes** | **Yes** | **Yes** | **7** |
| Uysal et al 2005 | **Yes** | **Yes** | **Yes** | **Yes** | **NA** | **Yes** | **Yes** | **Yes** | **7** |
| Innocenti et al 2005 | **Yes** | **Yes** | **Yes** | **Yes** | **NA** | **Yes** | **Yes** | **Yes** | **7** |
| Ceylan and Gencer 2005 | **Yes** | **Yes** | **Yes** | **Yes** | **NA** | **Yes** | **Yes** | **Yes** | **7** |
| Bas et al 2005 | **Yes** | **Yes** | **Yes** | **Yes** | **NA** | **Yes** | **Yes** | **Yes** | **7** |
| Akritidis et al 2005 | **Yes** | **Yes** | **Yes** | **Yes** | **NA** | **Yes** | **Yes** | **Yes** | **7** |
| Yen et al 2003 | **Yes** | **Yes** | **Yes** | **Yes** | **NA** | **Yes** | **Yes** | **Yes** | **7** |
| Gasparetto et al 2003 | **Yes** | **Yes** | **Yes** | **Yes** | **NA** | **Yes** | **Yes** | **Yes** | **7** |
| Liu et al 2002 | **Yes** | **Yes** | **Yes** | **Yes** | **NA** | **Yes** | **Yes** | **Yes** | **7** |
| Janner et al 2000 | **Yes** | **Yes** | **Yes** | **Yes** | **NA** | **Yes** | **Yes** | **Yes** | **7** |
| Shinmura et al 1999 | **Yes** | **No** | **Yes** | **Yes** | **NA** | **NA** | **Yes** | **NA** | **5** |
| Nishikage et al 1999 | **Yes** | **No** | **Yes** | **Yes** | **NA** | **NA** | **Yes** | **NA** | **5** |
| Jackman 1999 | **Yes** | **Yes** | **Yes** | **Yes** | **NA** | **Yes** | **Yes** | **Yes** | **7** |
| Muin and Zurin 1998 | **Yes** | **Yes** | **Yes** | **Yes** | **NA** | **Yes** | **Yes** | **Yes** | **7** |
|  | **Yes** | **Yes** | **Yes** | **Yes** | **NA** | **Yes** | **Yes** | **Yes** | **7** |
| Crump et al 1998 | **Yes** | **Yes** | **Yes** | **Yes** | **NA** | **Yes** | **Yes** | **Yes** | **7** |
| Reiser et al 1997 | **Yes** | **Yes** | **Yes** | **Yes** | **NA** | **Yes** | **Yes** | **Yes** | **7** |
| Ajay et al 1996 | **Yes** | **Yes** | **Yes** | **Yes** | **NA** | **Yes** | **Yes** | **Yes** | **7** |
| Ghosh and Senapati 1994 | **Yes** | **Yes** | **Yes** | **Yes** | **NA** | **Yes** | **Yes** | **Yes** | **7** |
| Geissmann and Minamoto 1994 | **Yes** | **Yes** | **Yes** | **Yes** | **NA** | **Yes** | **Yes** | **Yes** | **7** |
| Cilow et al 1994 | **Yes** | **Yes** | **Yes** | **Yes** | **NA** | **Yes** | **Yes** | **Yes** | **7** |
| Afghani and Lieberman 1994 | **Yes** | **Yes** | **Yes** | **Yes** | **NA** | **Yes** | **Yes** | **Yes** | **7** |
| Shen et al 1993 | **Yes** | **Yes** | **Yes** | **Yes** | **NA** | **Yes** | **Yes** | **Yes** | **7** |
| Gee et al 1992 | **Yes** | **Yes** | **Yes** | **Yes** | **NA** | **Yes** | **Yes** | **NA** | **6** |
| Shibolet et al 1979 | **Yes** | **Yes** | **Yes** | **Yes** | **NA** | **Yes** | **Yes** | **Yes** | **7** |
| Lieberman et al 1970 | **Yes** | **Yes** | **Yes** | **Yes** | **NA** | **Yes** | **Yes** | **Yes** | **7** |
| Case records of the Massachusetts General Hospital 1953 | **Yes** | **Yes** | **Yes** | **Yes** | **NA** | **Yes** | **Yes** | **NA** | **6** |
| Tooke 1935 | **Yes** | **Yes** | **Yes** | **Yes** | **NA** | **Yes** | **Yes** | **Yes** | **7** |
| Rodin and Dickey 1928 | **Yes** | **Yes** | **Yes** | **Yes** | **NA** | **Yes** | **Yes** | **Yes** | **7** |

**Tool for evaluating the methodological quality of case reports and case series**

**Domains Leading explanatory questions**

Selection 1. Does the patient(s) represent(s) the whole experience of the investigator or is the selection method unclear to the extent that other patients with similar presentation may not have been reported?

Ascertainment 2. Was the exposure adequately ascertained?

3. Was the outcome adequately ascertained?

Causality

4. Were other alternative causes that may explain the observation ruled out?

5. Was there a challenge/re-challenge phenomenon?

6. Was there a dose–response effect?

7. Was follow-up long enough for outcomes to occur?

Reporting

8. Is the case(s) described with sufficient details to allow other investigators to replicate the research or to allow practitioners make

inferences related to their own practice?

4,5= 14
